# Cardiovascular risk prediction using metabolomic biomarkers and polygenic risk scores: A cohort study and modelling analyses

**DOI:** 10.1101/2023.10.31.23297859

**Authors:** Scott C. Ritchie, Xilin Jiang, Lisa Pennells, Yu Xu, Claire Coffey, Yang Liu, Joel T. Gibson, Praveen Surendran, Savita Karthikeyan, Samuel A. Lambert, John Danesh, Adam S. Butterworth, Angela Wood, Stephen Kaptoge, Emanuele Di Angelantonio, Michael Inouye

## Abstract

**Background and Aims:** Metabolomic biomarker scores and polygenic risk scores (PRS) have shown promise for improving cardiovascular disease (CVD) prediction, but have not yet been evaluated in the context of current prediction models (SCORE2) and ESC recommendations for 10-year prediction of fatal and non-fatal CVD.

**Methods:** Metabolomics biomarker scores were constructed and compared to PRS and SCORE2 in 297,463 UK Biobank participants (8,919 incident CVD cases) aged 40–69 without previous CVD, diabetes, or lipid-lowering treatment. Improvement in risk discrimination when added to SCORE2 was assessed using Harrel’s C-index. Improvement in risk stratification following ESC guideline risk thresholds was assessed using categorical net reclassification. Population modelling was subsequently applied to estimate the impact on CVD prevention if applied at scale.

**Results:** Risk discrimination provided by SCORE2 (C-index: 0.719) was similarly improved by addition of metabolomic biomarker scores (ΔC-index: 0.010 [0.009–0.012]) and PRSs (ΔC-index 0.009; [0.008–0.011]). Addition of both metabolomic biomarker scores and PRSs to SCORE2 yielded the largest improvement risk discrimination, with ΔC-index 0.018 (0.016–0.020). Concomitant improvements in risk stratification were observed in categorical net reclassification index, with net case reclassification of 11.99% (10.98–12.99%). Modelling metabolic biomarker scores and PRSs for targeted risk-reclassification increased the number of CVD events prevented per 100,000 screened from 209 to 368 (ΔCVD_prevented_: 160 [151–169]) while essentially maintaining the number of statins prescribed per CVD event prevented.

**Conclusions:** Combining NMR scores and PRSs with SCORE2 enhanced prediction of first-onset CVD and could have substantial population health benefit if applied at scale.

## Introduction

Circulating biomarkers play a central role in cardiovascular disease (CVD) risk scores recommended by clinical guidelines to identify high-risk individuals for primary CVD prevention^1–3^. Total cholesterol and high-density lipoprotein (HDL) cholesterol are routinely measured and used alongside demographic and lifestyle risk factors to assess 10-year risk of CVD using risk scores such as SCORE2^4^. However, clinical guidelines have long-recognised the need to identify new risk factors to improve primary prevention^1–3^. Depending on the risk score and treatment threshold, it is estimated that between 40–86% of first onset CVD events occur in otherwise apparently healthy adults who would be classified as low-risk using conventional risk factors^5^. Efforts to improve CVD risk prediction models have considered additional circulating biomarkers^6^, such as C-reactive protein (CRP)^7,8^, as well as incorporating polygenic risk scores (PRSs) to account for genetic predisposition^9–11^. While PRSs have shown potential to enhance CVD risk screening^12–15^, addition of individual CVD biomarkers have thus far shown limited overall incremental benefits^16–18^.

High-throughput nuclear magnetic resonance (NMR) spectroscopy has enabled rapid and simultaneous quantification of several biomarkers from a single human blood plasma sample^19,20^. These include cholesterols and other lipids in lipoprotein sub-fractions, fatty acids, ketone bodies, amino acids, glycolysis metabolites and inflammation. NMR metabolic biomarker data has been quantified in numerous cohorts over the last decade, helping derive new insights into the genetic determinants, molecular pathogenesis, and epidemiology of CVD^21^. Several studies have investigated the utility of biomarkers combinations from NMR platforms to improve prediction of first-onset CVD^22–25^; however, they have focused on multi-disease prediction, used outdated clinical risk prediction scores, and have not investigated improvements relative to clinically relevant guideline-recommended risk thresholds.

Here, we utilize NMR biomarker data in UK Biobank to assess whether NMR biomarkers, in isolation or combination (*i.e.*, NMR scores), can improve 10-year CVD risk prediction in apparently healthy adults when added to the SCORE2 risk model, which is recommended by the European Society of Cardiology (ESC) 2021 guidelines for primary prevention of CVD^3^. We further assess whether incremental improvements in CVD risk prediction are meaningful at ESC 2021 recommended risk thresholds for treatment consideration^3^. In addition we compared the improvement in risk prediction provided by NMR scores to that provided by PRS^12^ and also assessed the PRSs and NMR scores combined. Finally, we modelled the potential public health benefits for CVD prevention if applied to the UK primary care population according to the ESC 2021 guidelines for statin initiation. A schematic of the overall study is given in **Figure 1**.

**Figure 1:**
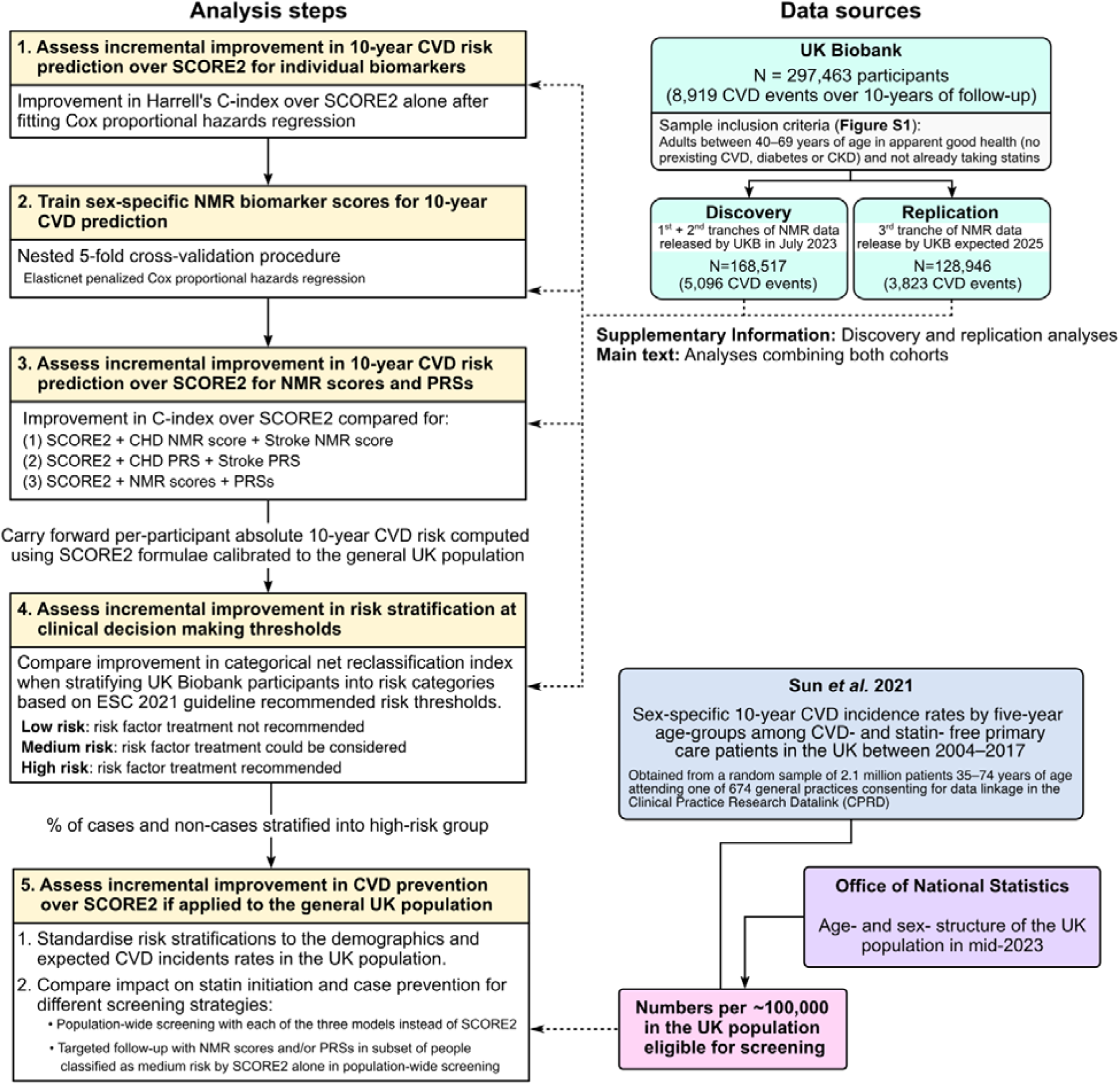
Study design. For details on sample inclusion and exclusion criteria when defining study eligibility for UK Biobank participants see **Figure S1**. Details on the NMR score training procedure are provided in the Supplementary Methods.

## Methods

### Study cohort

Modelling was performed in UK Biobank^26,27^ participants who were eligible for primary 10-year CVD risk assessment with SCORE2^3^: those who were 40–69 years of age at baseline assessment and had no prior history of established atherosclerotic cardiovascular disease, diabetes mellitus, chronic kidney disease, or familial hypercholesterolemia. Prevalent atherosclerotic cardiovascular disease included acute myocardial infarction, acute coronary syndromes, stroke, transient ischaemic attack, peripheral arterial disease, and history of revascularization procedures. Participants already taking lipid lowering medications were also excluded as the primary goal of the study was to improve identification of apparently healthy adults who would most benefit from lipid lowering medications for primary prevention of CVD. Participants were also excluded if they did not consent to electronic health record linkage, had incomplete data on SCORE2 risk factors, were ineligible for PRS assessment, or failed NMR biomarker quality control. Further information on the sample exclusion criteria is detailed in the **Supplementary Methods**. A flowchart showing the impact of each sample exclusion criteria on the cohort size and CVD cases is shown in **Figure S1**.

Of the 502,207 participants enrolled in UK Biobank consenting to electronic health record linkage, 297,463 participants met the inclusion criteria for this study (**Figure S1**). During the 2,897,497 person-years at risk (median [5^th^, 95^th^ percentile] follow-up of 10.0 [8.4–10.0] years), 8,919 CVD cases were recorded. Baseline cohort characteristics are detailed in **Table 1**.

**Table 1:**
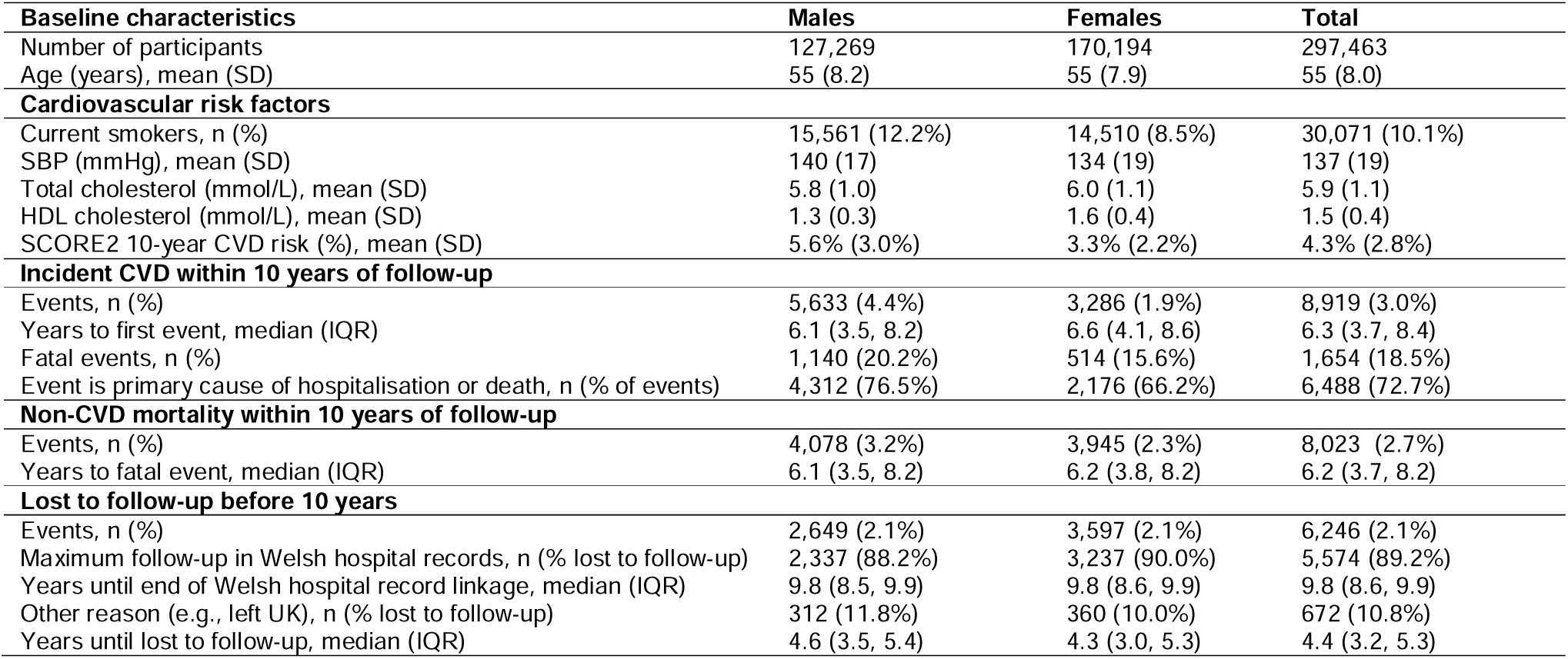
Cohort characteristics. SBP: systolic blood pressure. HDL: High density lipoprotein. LDL: Low density lipoprotein. sd: standard deviations. IQR: interquartile range.

| Baseline characteristics | Males | Females | Total |
| --- | --- | --- | --- |
| Number of participants | 127,269 | 170,194 | 297,463 |
| Age (years), mean (SD) | 55 (8.2) | 55 (7.9) | 55 (8.0) |
| **Cardiovascular risk factors** |  |  |  |
| Current smokers, n (%) | 15,561 (12.2%) | 14,510 (8.5%) | 30,071 (10.1%) |
| SBP (mmHg), mean (SD) | 140 (17) | 134 (19) | 137 (19) |
| Total cholesterol (mmol/L), mean (SD) | 5.8 (1.0) | 6.0 (1.1) | 5.9 (1.1) |
| HDL cholesterol (mmol/L), mean (SD) | 1.3 (0.3) | 1.6 (0.4) | 1.5 (0.4) |
| SCORE2 10-year CVD risk (%), mean (SD) | 5.6% (3.0%) | 3.3% (2.2%) | 4.3% (2.8%) |
| **Incident CVD within 10 years of follow-up** |  |  |  |
| Events, n (%) | 5,633 (4.4%) | 3,286 (1.9%) | 8,919 (3.0%) |
| Years to first event, median (IQR) | 6.1 (3.5, 8.2) | 6.6 (4.1, 8.6) | 6.3 (3.7, 8.4) |
| Fatal events, n (%) | 1,140 (20.2%) | 514 (15.6%) | 1,654 (18.5%) |
| Event is primary cause of hospitalisation or death, n (% of events) | 4,312 (76.5%) | 2,176 (66.2%) | 6,488 (72.7%) |
| **Non-CVD mortality within 10 years of follow-up** |  |  |  |
| Events, n (%) | 4,078 (3.2%) | 3,945 (2.3%) | 8,023 (2.7%) |
| Years to fatal event, median (IQR) | 6.1 (3.5, 8.2) | 6.2 (3.8, 8.2) | 6.2 (3.7, 8.2) |
| **Lost to follow-up before 10 years** |  |  |  |
| Events, n (%) | 2,649 (2.1%) | 3,597 (2.1%) | 6,246 (2.1%) |
| Maximum follow-up in Welsh hospital records, n (% lost to follow-up) | 2,337 (88.2%) | 3,237 (90.0%) | 5,574 (89.2%) |
| Years until end of Welsh hospital record linkage, median (IQR) | 9.8 (8.5, 9.9) | 9.8 (8.6, 9.9) | 9.8 (8.6, 9.9) |
| Other reason (e.g., left UK), n (% lost to follow-up) | 312 (11.8%) | 360 (10.0%) | 672 (10.8%) |
| Years until lost to follow-up, median (IQR) | 4.6 (3.5, 5.4) | 4.3 (3.0, 5.3) | 4.4 (3.2, 5.3) |

### Cardiovascular disease endpoint

The primary endpoint of the study was first-onset CVD within 10-years. CVD events were extracted from linked hospital episode statistics and death registry data (**Supplementary Methods**) following the definition used by the SCORE2 working group and ESC Cardiovascular risk collaboration^4^. This included International Classification of Diseases (ICD) codes covering: fatal hypertensive disease (ICD-10 codes I10–I16), fatal ischaemic heart disease (ICD-10 codes I20–I25), fatal arrhythmias or heart failure (ICD-10 codes I46–I52, excluding I51.4), fatal cerebrovascular disease (ICD-10 codes I60–I69, excluding I60, I62, I67.1, I68.2, and I67.1), fatal atherosclerosis or abdominal aortic aneurysm (ICD-10 codes I70–I73), sudden death and death within 24 hours of symptom onset (ICD-10 codes R96.0 and R96.1), non-fatal myocardial infarction (ICD-10 I21–I23), and non-fatal stroke (ICD-10 codes I60–I69, excluding I60, I62, I67.1, I68.2, and I67.1).

### Study Design

Study participants were further split into a discovery and replication cohort for all analyses based on the timing of the availability of NMR metabolomics data (**Figure 1, Figure S1**). The discovery cohort comprised the subset of 167,517 eligible study participants (5,096 CVD cases) for whom NMR data was made publicly available by UK Biobank in July 2023. The replication cohort comprised the remaining 128,946 participants (3,823 CVD cases) for whom we had early access to their NMR metabolomics data under UK Biobank project #30418 from Q3 of 2024 onwards. Allocation of participants to measurement tranches was randomised by UK Biobank with respect to phenotypes. Characteristics of the discovery and replication cohorts were similar (**Table S1**). Results presented in the main text are from pooled analyses across the full study cohort, with key differences from discovery and replication cohorts noted. Results in the discovery and replication cohorts for all analyses are presented in the corresponding supplementary figures and tables.

### Incremental value in 10-year CVD risk prediction for individual biomarkers

Improvements in 10-year CVD risk discrimination were assessed by changes in Harrell’s C-index (ΔC-index) beyond SCORE2 alone. SCORE2 was computed from age, sex, smoking status, systolic blood pressure (SBP), total cholesterol, and high-density lipoprotein (HDL) cholesterol using established coefficients^4^. The total and HDL cholesterol measures used were those quantified by clinical biochemistry assays (Beckman Coulter Inc)^28^. Further details on data quantification, SCORE2 computation, and statistical modelling are provided in the **Supplementary Methods**.

Improvements in 10-year CVD risk discrimination were assessed for each of the 249 NMR biomarkers^29,30^ (**Table S2**) and 28 clinical chemistry biomarkers^28^ (**Table S3**). In the discovery cohort, sex-stratified Cox proportional hazards regressions were fit for 10-year CVD risk for each biomarker, with the biomarker as an independent variable and SCORE2 as an offset term. In the replication cohort, the model fit for each biomarker was predicted using hazard ratios obtained from the discovery cohort. Predicted model fits were pooled across the discovery and replication cohorts for analyses presented in the main text. P-values were corrected for multiple testing across the 277 tested biomarkers using Benjamini-Hochberg false-discovery rate (FDR) correction. Further details are given in the **Supplementary Methods**.

### Incremental value in 10-year CVD risk prediction for biomarker combinations and PRSs

To assess whether combinations of biomarkers could improve 10-year CVD risk discrimination we assessed (1) a model combining SCORE2 with NMR biomarker scores trained using machine learning models to maximise CVD prediction (**Supplementary Methods**), and (2) a multivariable model of SCORE2 + clinical biochemistry biomarkers passing FDR significance in assessment of individual biomarkers above. NMR biomarkers and clinical chemistry biomarkers were modelled separately and with differing approaches as the 249 NMR biomarkers can be obtained simultaneously from a single measurement^19^ and aggregation of their disease associations into biomarker scores has shown promising potential in risk prediction settings^22–25^. In contrast, the clinical chemistry biomarkers represent a collection of expert-selected biomarkers that cannot be simultaneously obtained from a single measurement: requiring multiple biochemistry platforms and providers each requiring different sample preparation techniques^28^.

Models combining SCORE2 with NMR biomarker scores or clinical chemistry biomarkers were additionally compared to and combined with PRSs for coronary heart disease^9^ (Polygenic Score Catalog^31^ accession PGS000018) and ischaemic stroke^10^ (PGS000039) which have previously been shown to enhance 10-year CVD risk prediction when added alongside conventional risk factors^12^. Notably, PRSs capture inherited lifetime risk due to genetics^9–11^, whereas biomarkers capture part of the dynamic component of risk conferred by lifestyle and environment^22^.

In total, we compared five models to SCORE2: (1) SCORE2 + clinical biomarkers, (2) SCORE2 + NMR scores, (3) SCORE2 + PRSs, (4) SCORE2 + clinical biomarkers + PRSs, and (5) SCORE2 + NMR scores + PRSs. Further details on the multivariable model fits are given in the **Supplementary Methods**. Improvements in 10-year CVD risk discrimination were assessed as described above, with multivariable models fit in the discovery cohort and predicted in the replication cohort.

### Incremental value in 10-year CVD risk stratification at risk thresholds used for clinical decision making

Categorical net reclassification improvement (NRI) analysis^32,33^ was used to assess the incremental value of the five multivariable models over SCORE2 for stratifying individuals based on ESC 2021 recommended risk thresholds for treatment consideration^3^. For each model, study participants <50 years of age were stratified into low, medium, and high–risk groups if their predicted absolute risk was <2.5%, <7.5%, and ≥7.5% respectively, and study participants ≥50 years of age were stratified at <5%, <10%, ≥10% respectively. Categorical NRI analysis was used to assess (1) the % of incident CVD cases correctly reclassified from a lower risk group into higher risk group, and (2) the % of non-cases correctly reclassified from a higher risk group into a lower risk group. Further details on the computation of absolute risks for each model and the NRI analysis are provided in the **Supplementary Methods**.

### Incremental value for CVD prevention for population-wide and targeted screening

Two different screening strategies were assessed for incremental benefits for primary prevention of CVD if applied to the UK primary care population eligible for screening: (1) population-wide screening, in which all people eligible for screening were assessed with each model; and (2) targeted screening, in which people were first stratified into low, medium, and high-risk groups using SCORE2 alone, then those allocated to the medium risk group were re-assessed using the models adding NMR scores, clinical biomarkers, and/or PRSs to SCORE2.

Incremental improvements in primary CVD prevention for each alternative model were assessed by differences from SCORE2 alone per 100,000 screened in (1) the number classified as high risk (ΔN_high-risk_); (2) the number of future CVD cases amongst the high-risk group (ΔCVD_high-risk_); (3) the number of future CVD events expected to be prevented by initiation of statins in the high-risk group (ΔCVD_prevented_); (4) the number needed to screen to prevent one CVD event (ΔNNS); and the number of statins prescribed per CVD event prevented (ΔNNT). The impact of statin initiation was modelled as preventing one in five incident CVD events^34^.

To account for the healthy ascertainment bias of UK Biobank^26^, numbers of CVD cases and non-cases stratified into each risk group by each model were standardised to the demographics and expected CVD incidents rates of the UK primary care population eligible for screening. Demographic data on age and sex distributions of the general UK population were obtained from mid-2023 estimates published by the Office for National Statistics^35^ and 10-year CVD incident rates were obtained from previously published estimates^12^ extracted from CVD- and statin-free primary care patients. Standard errors, 95% confidence intervals, and P-values were obtained in a bootstrap procedure with 1,000 bootstraps. Further details are provided in the **Supplementary Methods**.

### Sensitivity analyses

Throughout the study we compared results to those obtained from analyses performed in males and females separately, using QRISK3 instead of SCORE2 as the risk score^36^, and using narrower set of criteria for identifying incident CVD events. The more narrowly defined CVD endpoint was defined to include only incident myocardial infarction (ICD-10 codes I21 and I22), fatal coronary heart disease (ICD-10 codes I20– I25), and fatal cerebrovascular events (ICD-10 codes I60–I69 or F01). Further details on QRISK3 and its computation are given in the **Supplementary Methods**.

### Ethics statement

This study was approved under UK Biobank Projects 30418 and ethics approval was obtained from the North West Multi-Center Research Ethics Committee. UK Biobank participants provided written informed consent for health related research^26^.

### Data Availability

All data described are available through UK Biobank subject to approval from the UK Biobank access committee. See https://www.ukbiobank.ac.uk/enable-your-research/apply-for-access for further details.

### Code Availability

Code underlying this paper are available at https://github.com/sritchie73/UKB_NMR_CVD_prediction/. This repository and specific release for this paper are permanently archived by Zenodo at https://doi.org/10.5281/zenodo.15475143.

## Results

### Incremental value in 10-year CVD risk prediction for individual biomarkers

Across all UK Biobank participants eligible for CVD screening for primary prevention (n=297,463) the sex-stratified C-index for SCORE2 alone was 0.719 (95% confidence interval [CI]: 0.714, 0.723) for predicting first-onset CVD within 10 years of follow-up (8,919 cases). When testing improvements in CVD risk discrimination for each of the 249 NMR biomarkers (**Table S2**) and 28 clinical chemistry biomarkers individually (**Table S3**) we observed statistically significant improvement in C-index for 66 of the 277 biomarkers (**Table S4**) based on a false discovery rate (FDR) adjusted P-value < 0.05. Improvements in sex-stratified C-index over SCORE2 (ΔC-index) were observed (**Figure 2A**). The largest ΔC-index observed for any biomarker was with addition of cystatin-C measured by clinical biochemistry assay (**Figure 2A**), with ΔC-index of 0.006 (95% CI: 0.005, 0.007; P-value: 2×10^−29^; FDR: 6×10^−27^; **Table S4**). The largest ΔC-index observed for any of the 249 NMR biomarkers was with addition of albumin (**Figure 2A**), with ΔC-index of 0.005 (95% CI: 0.004, 0.006; P-value: 4×10^−16^; FDR: 4×10^−14^; **Table S4**).

**Figure 2:**
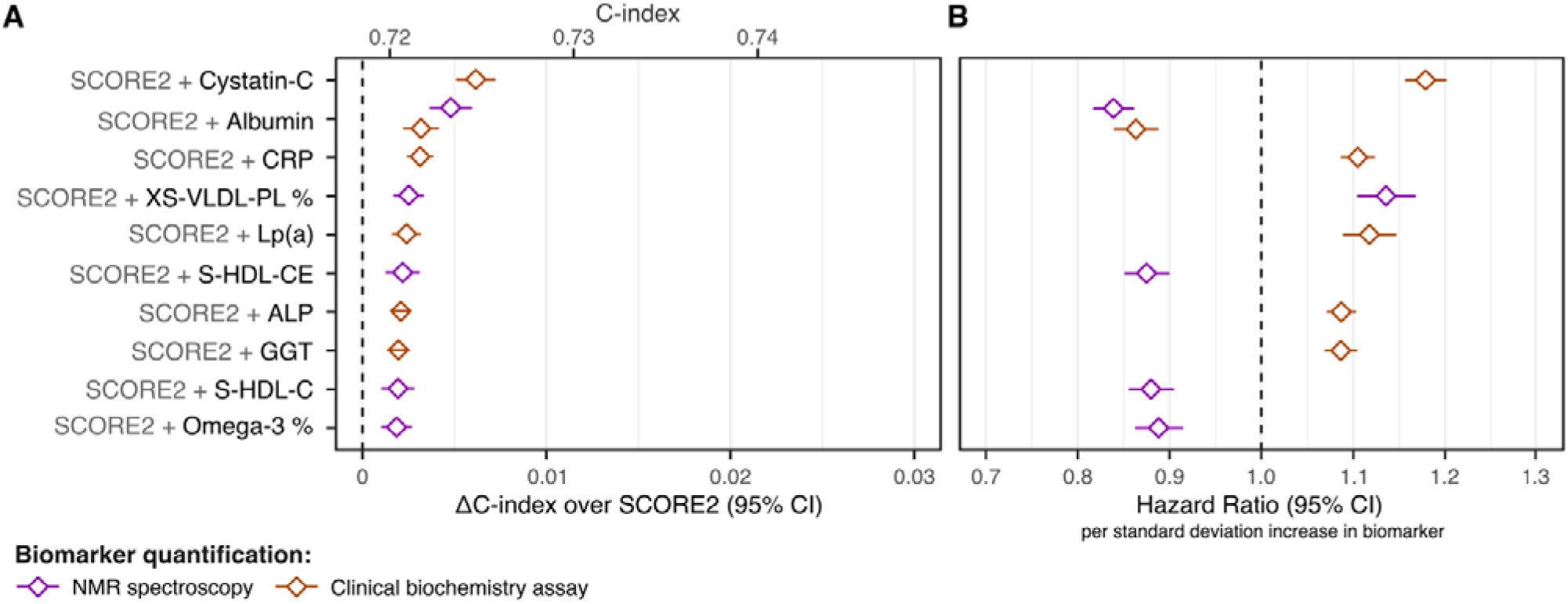
Incremental 10-year CVD risk discrimination over SCORE2 for individual biomarkers. **A)** Change in C-index (ΔC-index) relative to SCORE2 for the top 10 biomarkers by ΔC-index. The ΔC-index were assessed for each biomarker and quantification method separately (277 tests total; **Table S4**) in the subset of the 297,463 study participants (8,919 CVD cases) with non-missing biomarker concentrations for each test. Among the top 10 biomarker shown, only albumin concentrations were quantified by both clinical biochemistry assays (orange) and high-throughput NMR spectroscopy (purple). **B)** Hazard ratios per standard deviation increase in the respective biomarker concentration in the sex-stratified Cox proportional hazards model fit with SCORE2 as an offset term in the 168,517 study participants in the discovery cohort, which were used to predict the SCORE2 + biomarker model fit in the replication cohort (**Methods**).

Results were similar when analysing the discovery (n=167,517; 5,096 CVD cases) and replication (n=128,946; 3,823 CVD cases) cohorts separately (**Figure S2A**, **Table S5**). Results were also similar when analysing males (n=127,269; 5,633 CVD cases) and females (n=170,194; 3,286 CVD cases) separately, with ΔC-index estimates consistent for the strongest biomarkers (**Table S2B**, **Table S5**). Results were also similar when using a more narrowly defined CVD definition (**Figure S2C**, **Table S5**). Improvements in ΔC-index from individual biomarkers were smaller when using QRISK3 instead of SCORE2 as the conventional risk score (**Figure S2D**) with 16 of 277 biomarkers significantly increasing C-index at FDR < 0.05 (**Table S5**).

### Incremental value in 10-year CVD risk prediction for biomarker combinations and PRSs

When adding NMR biomarker scores (**Table S6**, **Supplementary Methods**) or the 11 FDR-significant clinical biochemistry biomarkers to SCORE2 (**Figure 3**, **Table S7**) we observed statistically significant improvements in ΔC-index of 0.010 (95% CI: 0.009, 0.012; P-value: 1×10^−32^) and 0.014 (95% CI: 0.012, 0.015; P-value: 7×10^−47^) respectively (**Figure 4A, Table S8**). Addition of PRSs to SCORE2 yielded a similar improvement in ΔC-index of 0.009 (95% CI: 0.008, 0.011; P-value: 1×10^−30^). Improvement in risk discrimination was greatest when combining multiple clinical biomarkers or NMR scores with PRSs (**Figure 4A**), with ΔC-index of 0.021 (95% CI: 0.019, 0.023; P-value: 3×10^−74^) and ΔC-index of 0.018 (95% CI: 0.016, 0.020; P-value: 4×10^−58^) respectively (**Table S8**)—an 9.6% gain in C-index relative to SCORE2 alone—for a total absolute C-index of 0.740 compared to 0.719 for SCORE2 alone.

**Figure 3:**
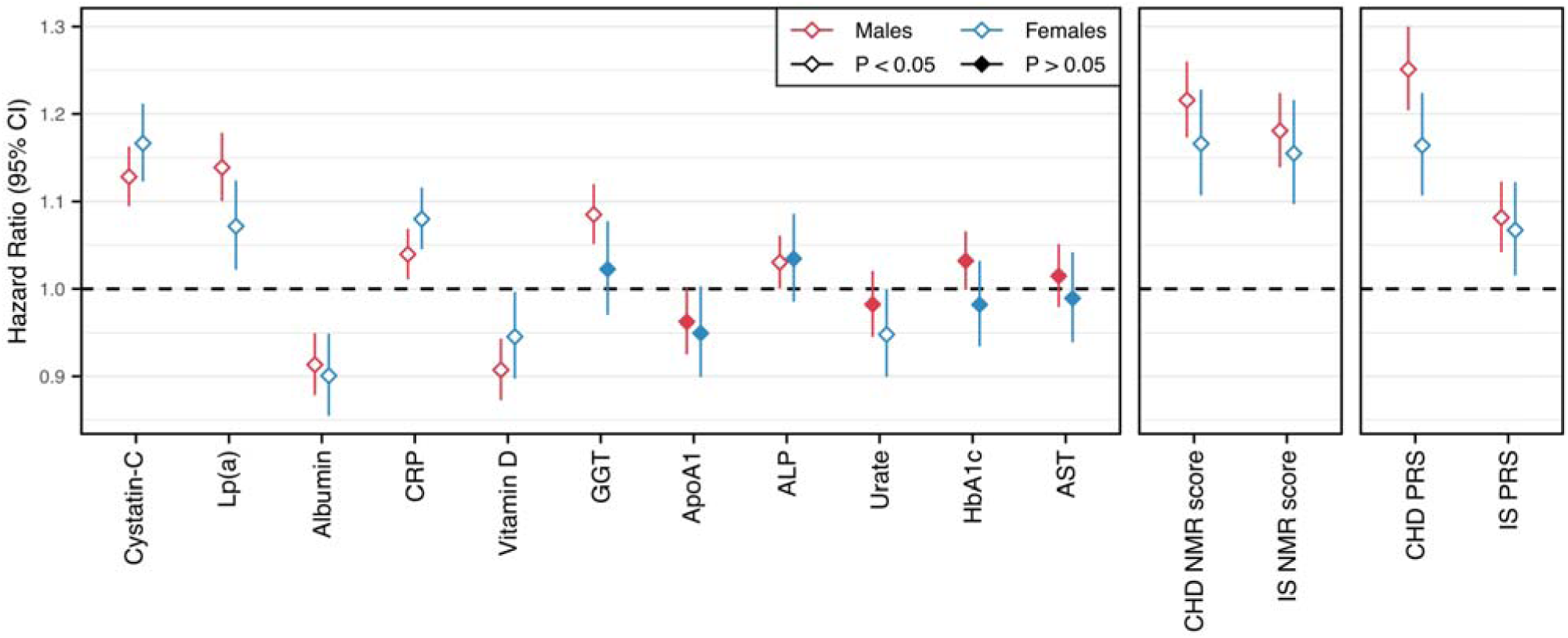
Multivariable model fits for combining biomarkers, NMR scores, and PRSs with SCORE2. Sex-specific Cox proportional hazards regressions with SCORE2 as an offset term fit in the discovery cohort (n=168,517; 5,096 CVD cases) to create multivariable models of (1) SCORE2 + clinical biomarkers (2) SCORE2 + NMR scores, and (3) SCORE2 + PRSs. Hazard ratios are per standard deviation increase in the respective biomarker, NMR score, or PRS and are detailed in **Table S7**. Log hazard ratios were used to predict the sex-specific models in the replication cohort (**Supplementary Methods**). The 11 clinical biomarkers used were those that had FDR significant ΔC-index over SCORE2 alone when testing individual biomarkers (**Table S4**). Details on the NMR scores are given in **Table S6** and the **Supplementary Methods**. CHD: coronary heart disease. IS: ischaemic stroke. Lp(a): Lipoprotein(a). CRP: C-reactive protein. GGT: Gamma glutamyltransferase. ApoA1: Apolipoprotein A1. ALP: Alkaline phosphatase. HbA1c: Glycated haemoglobin. AST: aspartate aminotransferase.

**Figure 4:**
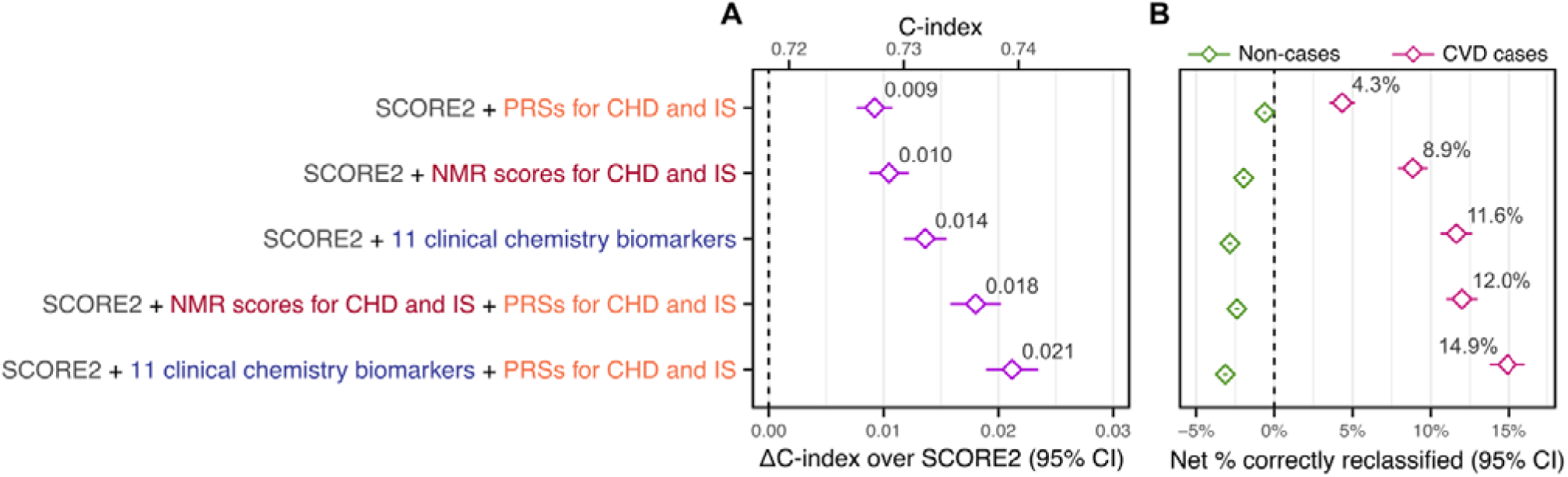
Incremental value in 10-year CVD risk prediction for biomarker combinations and PRSs. **A)** Incremental improvement in 10-year CVD risk discrimination over SCORE2 alone (ΔC-index) in the 297,463 study participants (8,919 CVD cases) when adding NMR scores, clinical biomarkers, and/or PRSs to SCORE2. NMR scores for coronary heart disease (CHD) and ischaemic stroke (IS) each comprised the 106 non-derived biomarkers on the NMR platform and are detailed in **Table S6** and the **Supplemen ary Methods**. The 11 clinical biochemistry biomarkers added to SCORE2 were cystatin-C, C-reactive protein, alkaline phosphatase, albumin, gamma glutamyltransferase, lipoprotein(a), aspartate aminotransferase, glycated haemoglobin (HbA1c), urate, vitamin D, and Apolipoprotein A1 (**Table S7**). The PRSs for CHD and IS added to SCORE2 were those published in the PGS Catalog with accession numbers PGS000018 and PGS000039 respectively. Details on the multivariable fits for each of the five models are provided in **Table S7** and the **Supplementary Methods**. C-indices, ΔC-indices, and 95% confidence intervals for each model are provided in **Table S8**. **B)** Categorical net reclassification improvement (NRI) index relative to SCORE2 when stratifying the 297,463 study participants (8,919 CVD cases) into low-, medium-, and high-risk categories based on the absolute 10-year CVD risk predicted by each model and ESC 2021 recommended risk thresholds for treatment consideration (**Methods**). Net % correctly reclassified: net % of CVD cases that were correctly reclassified into a higher risk category (pink) or net % of non-cases that were correctly reclassified into a lower risk category (green) when comparing the given model to SCORE2. Categorical NRI details are provided in **Table S10**. Numbers allocated to each risk category by each model are detailed in **Table S11**.

Results were similar when analysing the discovery and replication cohorts separately (**Figure S3A**, **Table S9**) with slightly higher ΔC-index estimates in the discovery cohort as expected. Results were also broadly similar when analysing males and females separately, with stronger improvements in ΔC-index in males than in females for models incorporating PRSs (**Figure S3B, Table S9**). Results were also similar when using a more narrowly defined CVD definition (**Figure S3C**, **Table S9**) and when using QRISK3 instead of SCORE2 as the conventional risk score (**Figure S3D**, **Table S9**). Notably attenuation of ΔC-index when using QRISK3 was less pronounced when combining information across multiple biomarkers (**Figure S3D**) compared to the attenuation observed when assessing biomarkers separately (**Figure S2D**).

### Incremental value in 10-year CVD risk stratification at risk thresholds used for clinical decision making

Statistically significant improvement in risk stratification over SCORE2 among incident CVD cases was observed for all five alternative models tested (**Figure 4B**, **Table S10–S11**), when using ESC 2021 recommended risk thresholds for treatment consideration^3^. Improvements in case classification when adding either the NMR biomarker scores or the 11 clinical biochemistry biomarkers to SCORE2 were more than twice as strong as those from PRSs. We observed a net case reclassification rate of 8.85% (95% CI: 7.90%, 9.80%; P-value: 8×10^−74^) with addition of NMR scores and 11.63% (95% CI: 10.62%, 12.64%; P-value: 5×10^−112^) with addition of the 11 clinical biomarkers, compared to 4.33% (95% CI: 3.52%, 5.15%; P-value: 2×10^−25^) with addition of PRSs (**Table S10**). Improvements in case classification were strongest when combining multiple clinical biomarkers or NMR scores with PRSs (**Figure 4B**), with a net case reclassification rate of 14.93% (95% CI: 13.77%, 16.08%; P-value: 2×10^−141^) and 11.99% (95% CI: 10.98%, 12.99%; P-value: 2×10^−121^) respectively (**Table S10**).

A statistically significant, inappropriate reclassification for non-cases was also observed for all five alternative models (**Figure 4B**, **Table S10**). The net reclassification rate for non-cases was −1.99% (95% CI: −2.16%, −1.81% ; P-value: 3×10^−199^) with addition of NMR scores, −2.84% (95% CI: −2.98%, −2.70%; P-value: <3×10^−308^) with addition of 11 clinical biomarkers, −0.61% (95% CI: −0.73%, −0.49%; P-value: 2×10^−23^) with the addition of PRSs, −2.39% (95% CI: −2.54%, −2.24%; P-value: 1×10^−218^) with addition of both NMR scores and PRSs, and −3.13% (95% CI: −3.28%, −2.98%; P-value: <3×10^−308^) with addition of both 11 clinical biomarkers and PRSs (**Table S10**).

Results were similar when analysing the discovery and replication cohorts separately (**Figure S4A**, **Table S12–S13**),when analysing males and females separately (**Figure S4B**), and when using a more narrowly defined CVD definition (**Figure S4C**). Notably, inappropriate net reclassification of non-cases in males was approximately half that of the inappropriate net reclassification of non-cases observed in females (**Figure S4B**, **Table S12**). Improvements in net case reclassification were strongly attenuated, although remained statistically significant, when adding NMR scores, 11 clinical biomarkers, and/or PRSs to QRISK3 and using the 10% risk threshold recommended alongside QRISK3 by the UK’s National Institute for Health and Care Excellence (NICE) 2023 guidelines for cardiovascular risk assessment and reduction^36^ (**Figure S4D**, **Table S12**). Notably, there was a strong increase in the proportion of participants classified as high risk by QRISK3 alone with increasing age (e.g. >99% of males >65 years of age; **Table S14**), suggesting the observed attenuation was a consequence of the dominating influence of age on treatment consideration when following the NICE 2023 guidelines with QRISK3.

### Incremental value for CVD prevention for population-wide and targeted screening

Consistent with the categorical NRI analyses above, we observed statistically significant incremental improvements in primary prevention of CVD when applying all five alternative models to the UK primary care population eligible for screening (**Figure 5A, Table S15**). For all five alternative models, we observed significant increases in N_high-risk_ and CVD_high-risk_, with concomitant increases in CVD_prevented_ and decreases in NNS. CVD_prevented_ per 100,000 screened increased from 208 with SCORE2 alone, to 254 with addition of PRSs (ΔCVD_prevented_: 47; 95% CI: 39, 55; PLvalue: 3×10^−32^), to 301 with addition of NMR scores (ΔCVD_prevented_: 92; 95% CI: 83, 101; PLvalue: 1×10^−94^), to 317 with addition of 11 clinical biomarkers (ΔCVD_prevented_: 118; 95% CI: 108, 128; PLvalue: 2×10^−125^), to 339 with addition of both NMR scores and PRSs (ΔCVD_prevented_: 130; 95% CI: 120, 141; PLvalue: 5×10^−138^), and to 356 with addition of 11 clinical biomarkers and PRSs (ΔCVD_prevented_: 157; 95% CI: 146, 168; PLvalue: 2×10^−169^) (**Table S15**). Importantly, our modelling indicated no statistically significant change in NNT, which was constant at 22 (**Figure 5A, Table S15**).

**Figure 5:**
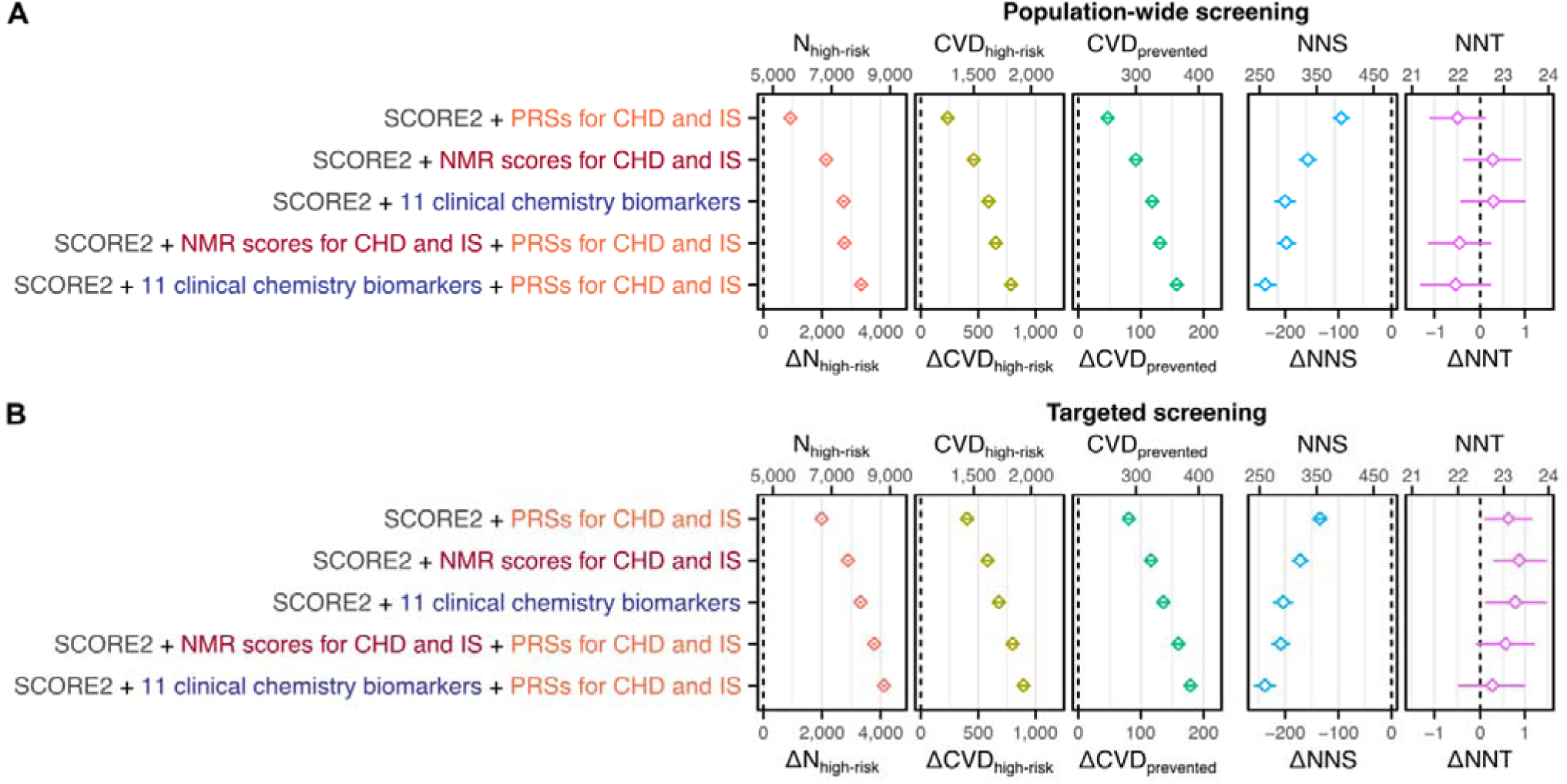
Incremental value for CVD prevention per 100,000 in the UK eligible for screening. **A)** Incremental value for primary prevention of CVD per 100,000 in the UK eligible for screening when adding NMR scores, clinical biomarkers, and/or PRSs to SCORE2. Per 100,000 the number of CVD events expected within 10 years was 6,686 (**Supplementary Methods**). **B)** Incremental value for primary prevention of CVD when using NMR scores, clinical biomarkers, and/or PRSs for targeted risk-reclassification in people classified as medium risk by SCORE2 alone (36,054 per 100,000; 3,765 CVD events). **A–B**) ΔN_high-risk_: change in the number per 100,000 classified as high risk relative to SCORE2 alone. ΔCVD_high-risk_: change in the number of incident CVD events classified as high-risk per 100 000 screened. ΔCVD_prevented_: change in the number of CVD events expected to be prevented over 10 years due to statin initiation in the high-risk group. ΔNNS: change in number needed to screen to prevent one CVD event. ΔNNT: change in the number of statins prescribed per CVD event prevented. Estimates were derived by standardising risk stratification in the study cohort to the demographics and expected CVD incidents rates of the UK primary care population eligible for screening (**Methods**). 95% confidence intervals were estimated via a bootstrap sampling procedure with 1000 bootstraps (**Methods**). Point estimates and 95% confidence intervals are detailed in **Table S15**.

Targeted re-screening of the population classified as medium risk by SCORE2 (36,054 per 100,000) with NMR scores, clinical biomarkers, and/or PRSs yielded slightly stronger incremental improvements in primary prevent of CVD (**Figure 5A, Table S15**). CVD_prevented_ per 100,000 screened increased from 208 with SCORE2 alone, to 288 with additional targeted screening with PRSs (ΔCVD_prevented_: 81; 95% CI: 74, 87; PLvalue: 8×10^−122^), to 325 with additional targeted screening with NMR scores (ΔCVD_prevented_: 116; 95% CI: 108, 124; PLvalue: 4×10^−186^), to 335 with additional targeted screening with 11 clinical biomarkers (ΔCVD_prevented_: 136; 95% CI: 127, 145; PLvalue: 3×10^−206^), to 368 with additional targeted screening with both NMR scores and PRSs (ΔCVD_prevented_: 160; 95% CI: 151, 169; PLvalue: 4×10^−257^), and to 378 with additional targeted screening with 11 clinical biomarkers and PRSs (ΔCVD_prevented_: 179; 95% CI: 169, 189; PLvalue: 4×10^−255^) (**Table S15**). A statistically significant, but small, increase in NNT was observed with additional targeted screening with either NMR scores, the 11 clinical biomarkers, or PRSs (ΔNNT: 1), which was attenuated (P-value > 0.05) when combining PRSs with NMR scores or clinical biomarkers (**Figure 5B**, **Table S15**).

Results were similar when analysing the discovery and replication cohorts separately (**Figure S5**, **Table S16**) and when using a more narrowly defined CVD definition (**Figure S6**, **Table S16**). Results were also similar in both males and females, including expected differences in magnitudes arising from differences in CVD incidence (9,025 per 100,000 males screened vs. 4,447 per 100,000 females screened; **Figure S7**, **Table S16**). Improvements in primary prevention of CVD when using QRISK3 and NICE 2023 guideline recommended risk thresholds were smaller than those observed when using SCORE2 (**Figure S8**, **Table S16**) as expected due to the dominating influence of age on risk stratification observed above (**Table S14**). Notably, the total number of statins prescribed (*i.e.*, N_high-risk_) and NNT were significantly higher with QRISK3 (N_high-risk_: 28,036; NNT: 34) than SCORE2 (N_high-risk_: 4,671; NNT: 22).

## Discussion

Determining the added value of biomarkers beyond total and HDL cholesterol for 10-year CVD risk prediction is an area of interest for enhancing CVD prevention^3^. Here, we investigated whether 10-year CVD risk prediction in UK Biobank participants eligible for screening could be improved, in comparison to the currently recommended SCORE2^3,4^.

We found statistically significant improvements in 10-year CVD risk prediction from 66 of 277 biomarkers quantified either individually by clinical chemistry assays or simultaneously by plasma NMR spectroscopy. Combining NMR biomarkers into NMR scores almost doubled the gain in observed predictive performance (ΔC-index) as compared to any single NMR biomarker. NMR biomarker scores and PRSs offered largely orthogonal information and increased SCORE2 C-index to similar degrees.

When added to SCORE2, the clinical biomarkers yielding the strongest improvements in 10-year CVD risk prediction were Cystatin-C, Lipoprotein(a), Albumin, C-Reactive Protein, and Vitamin D. Each of these are well-known biomarkers of CVD risk^37–41^, and for C-Reactive Protein and Lipoprotein(a), biomarkers of long-standing interest for CVD risk prediction^3,16,42^. A key CVD risk pathway intersected by all five biomarkers is inflammation^43–47^, which is a well-studied target in CVD prediction and prevention research^48^. Likewise, the strongest contributors to the NMR scores were Albumin and glycoprotein acetyls (GlycA); an NMR signal quantifying the levels of multiple inflammatory proteins^49,50^ and a stronger biomarker of chronic inflammation than C-Reactive Protein^51^, which has been associated with CVD risk in multiple studies^52–54^.

Risk prediction models including NMR scores and/or PRSs also improved risk stratification of future CVD events when using risk thresholds recommended for clinical decision making by the ESC 2021 guidelines for CVD prevention. NMR scores improved net case reclassification to a greater extent than PRSs (8.85% vs 4.33%, respectively); however, when combined, NMR scores and PRSs improved net case reclassification by 11.99%. These results highlight the complementary nature of the information capture by PRSs and NMR scores. While PRSs capture the lifetime risks due to genetics^9–11^, NMR scores capture part of the dynamic component of risk conferred by lifestyle and environment^22^, which act on that genetic background.

When modelling the potential benefits for primary prevention of CVD in the wider UK population eligible for screening, we found adding NMR scores and/or PRSs to SCORE2 significantly increased the those would be recommended for statin initiation (following the ESC 2021 guidelines for risk factor treatment for CVD prevention^3^) and who would subsequently experience a CVD event. Importantly, the number of statins prescribed per CVD event prevented would stay constant. We estimated that adding NMR scores to SCORE2 would increase the number of CVD events prevented from 208 to 301 (per 100,000); adding PRSs to SCORE2 would increase the number of CVD events prevented to 254; and adding both NMR scores and PRSs to SCORE2 would increase the number of CVD events prevented to 339.

To increase its efficiency, we also modelled the potential benefits of targeted follow-up screening in those at medium risk, for whom the ESC 2021 guidelines suggest considering, but do not explicitly recommend, risk factor treatment^3^. We estimated that, per 100,000 screened, that targeted risk-reclassification with NMR scores would increase the number of CVD events prevented to 325; targeted risk-reclassification with PRSs would increase the number of CVD events prevented to 288; and targeted risk-reclassification with both NMR scores and PRSs would increase the number of CVD events prevented to 368. We also estimated that this targeted follow-up screening with both NMR scores and PRSs would essentially maintain the number statins prescribed per CVD event prevented.

Across all our analyses, incremental gains in CVD prediction, risk stratification, and screening efficacy were slightly large when combining information from 11 clinical chemistry assays compared to using NMR scores. However, this additional incremental gain came with a trade-off of increased complexity and potentially cost; requiring quantification via multiple different assay types, providers, and in some cases, sample preparation techniques^28^. In contrast, NMR scores required a single assay, which included the total and HDL cholesterol measures required for SCORE2^19^.

This study has several limitations. While we were able to replicate our results using an independent set of UK Biobank participants with NMR data, we were unable to assess generalizability across populations. Notably, different regions of Europe vary in CVD risk profiles^4^ and thus may have different distributions of some biomarkers. Further, our study comprised almost entirely (>95%) European ancestries and thus may not generalize to other ancestry groups which have different risk profiles^55^. Importantly, development of SCORE2 required information on risk factor distributions from a range of cohorts to calibrate risk factor distributions and their estimates for 10-year CVD risk^4^, which would also need to be the case for any extensions to SCORE2 with additional biomarkers or PRSs. Our estimates of incremental improvement for primary prevention of CVD relied on a demographic standardisation procedure with multiple assumptions and sources of competing biases. For example, incremental benefits may be underestimated due to the healthy ascertainment bias of UK Biobank^26^, but at the same time may be overestimated as expected 10-year CVD incidence rates were estimated in CVD- and statin-free primary care patients^12^, whereas the age- and sex-demographics were obtained from the general UK population^35^ and thus could not account for screening eligibility with increasing age. However, whilst these may impact the precise magnitude of benefits for primary prevention of CVD, they do not impact the relative incremental benefits when comparing models developed in this study to SCORE2 alone.

Overall, this study represents the largest population health assessment of metabolomic and genomic biomarkers for CVD to date. While our findings suggest that there are potential gains for CVD risk prediction and prevention, there are obvious challenges for validating clinical utility and potential implementation. Commercial providers of NMR biomarkers and PRSs exist, yet fidelity, scale, and cost frequently mean that real world benefits are less than those estimated in prospective cohort studies. Nevertheless, our results indicate that current technologies that can scale to populations (e.g. NMR metabolomics and genomics) have the capacity to improve CVD risk prediction. However, further studies are needed to evaluate the efficacy and cost-effectiveness of NMR scores and PRSs for improving 10-year CVD risk prediction in these settings. For the goal of primordial prevention, further studies are also needed to investigate the potential for circulating biomarkers for CVD risk prediction in younger adults^56^.

In conclusion, our results indicate that incorporating scores of NMR metabolomic biomarkers into 10-year CVD risk prediction could enhance prediction of first-onset CVD. We further add to the growing body of evidence that PRSs can be used to enhance CVD risk prediction over conventional risk factors^11,12^ and show that improvements in 10-year CVD risk prediction from PRSs are orthogonal to, and can be combined with, NMR scores. Applied at scale, integrating NMR scores alongside PRSs with SCORE2 may have population health benefit.

## Acknowledgements

The authors are grateful to UK Biobank for access to data to undertake this study (Projects #30418).

Nightingale Health Plc is acknowledged for early access to the UK Biobank NMR biomarker data and discussions regarding sources of experimental variation.

This work was performed using resources provided by the Cambridge Service for Data Driven Discovery (CSD3) operated by the University of Cambridge Research Computing Service (www.csd3.cam.ac.uk), provided by Dell EMC and Intel using Tier-2 funding from the Engineering and Physical Sciences Research Council (capital grant EP/P020259/1), and DiRAC funding from the Science and Technology Facilities Council (www.dirac.ac.uk).

This work was supported by core funding from the British Heart Foundation (BHF) (RG/18/13/33946: RG/F/23/110103), National Institute for Health and Care Research (NIHR) Cambridge Biomedical Research Centre (BRC-1215-20014: NIHR203312) [*], BHF Chair Award (CH/12/2/29428), Cambridge BHF Centre of Research Excellence (RE/24/130011, RE/18/1/34212) and by Health Data Research UK, which is funded by the UK Medical Research Council, Engineering and Physical Sciences Research Council, Economic and Social Research Council, Department of Health and Social Care (England), Chief Scientist Office of the Scottish Government Health and Social Care Directorates, Health and Social Care Research and Development Division (Welsh Government), Public Health Agency (Northern Ireland), BHF and the Wellcome Trust.

S.C.R. was funded by a BHF Cambridge Centre for Research Excellence fellowship (RE/24/130011). X.J. was funded by a Wellcome Trust Fellowship (227566/Z/23/Z). L.P. and P.S. were supported by a Rutherford Fund Fellowship from the Medical Research Council grant MR/S003746/1. Y.X. and M.I. were supported by the UK Economic and Social Research Council (ES/T013192/1). S.A.L. was supported by a Canadian Institutes of Health Research postdoctoral fellowship (MFE-171279). E.D.A. holds a NIHR Senior Investigator Award. J.D. holds a BHF Professorship and a NIHR Senior Investigator Award. M.I. is supported by the Munz Chair of Cardiovascular Prediction and Prevention and the NIHR Cambridge Biomedical Research Centre (NIHR203312).

The funders had no role in study design, data collection and analysis, decision to publish, or preparation of the manuscript.

*The views expressed are those of the authors and not necessarily those of the NIHR or the Department of Health and Social Care.

