## Supplementary Information for "Cardiovascular risk prediction using metabolomic biomarkers and polygenic risk scores: A cohort study and modelling analyses"

### Supplementary Methods

#### Study cohort

UK Biobank is a cohort comprised of ~500,000 participants 35–75 years of age with written informed consent for health related research^26,27^. Participants were members of the UK population recruited through primary care lists whom accepted invitation to attend one of 22 assessment centres across the UK between 2006 and 2010^26^.

In this study we analysed a subset of 297,463 participants who at baseline assessment (1) consented for electronic health record linkage, (2) were eligible for 10-year CVD risk prediction with SCORE2^3,4^, (3) were not prescribed statins or other lipid lowering medications, and (4) had completed information on risk factors required for SCORE2 computation, (5) had NMR biomarker data (with <5% missingness), (6) had imputed genotypes, and (7) were eligible for joint analysis with PRSs. **Figure S1** shows the sample exclusions at each step.

Study participants were further split into a discovery and replication cohort for all analyses based on the timing of the availability of NMR metabolomics data (**Figure 1, Figure S1**). The discovery cohort comprised the subset of 167,517 eligible study participants for whom NMR data was made publicly available by UK Biobank in July 2023. The replication cohort comprised the remaining 128,946 participants for whom we had early access to their NMR metabolomics data under UK Biobank project #30418 from Q3 of 2024 onwards. Allocation of participants to measurement tranches was randomised by UK Biobank with respect to phenotypes.

#### Sample exclusion criteria

Eligibility for 10-year CVD risk prediction with SCORE2 was determined following the ESC 2021 guidelines for CVD prevention in clinical practice^3^. UK Biobank participants were included if at baseline assessment they were (1) 40 years of age or older and less than 70 years of age, (2) did not have established atherosclerotic cardiovascular disease (ASCVD), (3) did not have diabetes mellitus, (4) did not have chronic kidney disease, and (5) did not have familial hypercholesterolemia.

Disease history was determined using a combination of self-reported medical history (UK Biobank fields #6150, #4728, #20002, and #20004), prescription medications (fields #6153, #6177, and #20003), and retrospective linkage to hospital episode statistics (fields #41259, #41234, and #41149). Prevalent ASCVD included acute myocardial infarction, acute coronary syndromes, transient ischaemic attack, peripheral arterial disease, and history of revascularization procedures. A full list of ICD codes and self-report codes used to define ASCVD are given in **Table S17**. Prevalent diabetes mellitus was determined using the Eastwood *et al.* algorithms^57^, and included participants with probable or possible type 1 or type 2 diabetes. Prevalent chronic kidney disease was determined using the UK Biobank algorithmically defined outcome for end-stage renal disease (field #42026).

Familial hypercholesterolemia (FH) was determined using the Dutch Lipid Clinical Network (DLCN) diagnostic criteria^58^ as described in the ESC 2021 guidelines for CVD prevention in clinical practice^3^. Participants were excluded where they had low density lipoprotein (LDL) cholesterol quantified by clinical biochemistry assay ≥ 8.5 mmol/L (8 points on the DLCN diagnostic score indicative of probable FH) (fields #30780 and #30786). All participants with possible clinical history used for the DLCN diagnostic criteria were excluded due to prevalent ASCVD. Physical examination and family history data relevant to the DLCN diagnostic criteria were not collected at UK Biobank assessment. Functional mutations in the *LDLR, APOB*, and *PCSK9* genes were not assessed in this study.

Participants already prescribed statins or other lipid lowering medications for CVD prevention were excluded as the primary purpose of 10-year CVD risk prediction with SCORE2 is to identify high-risk individuals for statin initiation in apparently healthy adults^3^. These participants were identified as those who at baseline assessment self-reported cholesterol lowering medication on the touchscreen questionnaire on health and medical history (field #6153 for women and field #6177 for men) and/or had been prescribed any of 18 lipid lowering prescription medications for CVD prevention (field #20003). The list of qualifying prescription medications (**Table S18**) was determined by cross-referencing the list of medications present in UK Biobank with the British National Formulary^59^ chapter 2.12: Lipid-regulating drugs with the restriction that the drug indications must include prevention of cardiovascular diseases.

Participants were also excluded where it was not possible to predict 10-year CVD risk with SCORE2 due to missing quantitative risk factor information. Quantitative risk factors with missing data included systolic blood pressure (SBP) (missing data at baseline for all instances of fields #93 and #4080) and total cholesterol and high-density lipoprotein (HDL) cholesterol as measured by clinical biochemistry assays (fields #30690 and #30760 respectively).

Participants with >5% missing NMR biomarker data were excluded, as after removal of technical variation, this excess missing data primarily arose due to removal of outlier plates of non-biological origin (89% of missing values).

Finally, participants were excluded if they were used as part of the training for the PRSs for CHD (PGS000018)^9^ and ischaemic stroke (PGS000039)^10^ analysed in this study.

#### Electronic health records

UK biobank participants were linked by UK Biobank to hospital inpatient admissions records (fields #41259, #41234, and #41149) for hospitals in England, Wales, and Scotland and to national death registry records (fields #40000, #40001, and #40002). All incident hospital events or death records were coded with ICD-10 codes. Surgical procedures were coded with Office of Population Census and Surveys (OPCS) codes, and all incident surgical procedures were coded with OPCS-4 codes. Hospital and death records follow-up was available up to 6^th^ March 2018 for events occurring in hospitals in Wales, and to 2021 (beyond 10 years of follow-up) for events occurring in hospitals in England and Scotland.

Retrospective follow-up in hospital records was available from 27^th^ July 1993 for events occurring in hospitals in England, 2^nd^ December 1980 for events occurring in hospitals in Scotland, and 18^th^ April 1991 for events occurring in hospitals in Wales, with median of 15.75 years retrospective follow-up (maximum 29.76 years). Retrospective hospital events were coded with a combination of ICD-10 and ICD-9 codes (or OPCS-4 and OPCS-3 codes for surgical procedures).

Follow-up time for each participant was restricted to a maximum of 10 years, defined as the difference in years between baseline assessment and the earliest of the following records: (1) the date of the first CVD event, (2) the date of death, (3) the date lost to follow-up (fields #190 and #191; e.g. participant reported to NHS or UK Biobank as having left the UK), (4) the maximum follow-up date in hospital records from Wales—6^th^ March 2018—for participants located in Wales at baseline assessment or inferred to have moved to Wales since baseline assessment (based on presence of hospital records from Wales before 2018 and none in England or Scotland after, or change in location for subsequent UK Biobank assessments), or (5) date of baseline assessment plus ten years. Cohort characteristics reported in **Table 1** include details on the number of people with non-CVD related mortality or otherwise lost to follow-up prior to 10 years.

Participants with withdrawn consent for electronic health record linkage were identified from field #190 for sample exclusion.

#### NMR biomarker data quantification and quality control

NMR metabolite biomarker data was quantified in all ~500,000 participants as previously described^19,29^. Briefly, NMR spectroscopy (Nightingale Health Plc.) was used to measure the absolute concentrations of 168 biomarkers and 81 biomarker ratios from non-fasting plasma samples (UK Biobank aliquot 3). Details on the identity of the 249 NMR metabolite biomarkers are provided in **Table S2**.

Technical variation was subsequently removed using a modified version of our previously described pipeline^30^ that has been updated to reflect our exploration the additional ~380,000 participants measured since the pipeline development. This updated pipeline is made available as part of the ukbnmr R package. Briefly, technical variation removed included (1) time between sample preparation and sample measurement, (2) systematic differences in biomarker concentrations in each shipping batch due to sample position on the 96-well shipment plate, (3) measurement drift over time, (4) inter-spectrometer differences, and (5) shipment plates with systematically extreme concentrations of non-biological origin. For further details on the pipeline and modifications made with each UK Biobank data release see https://github.com/sritchie73/ukbnmr. The pipeline made available in version 2.2.1 of the ukbnmr package was used to remove technical variation from the discovery cohort, while for the replication cohort version 3.2.0 was used.

#### Clinical biochemistry assay quantification and quality control

Targeted blood biochemistry assays were quantified in all ~500,000 participants as previously described^28^. Briefly, absolute concentrations of 30 circulating biomarkers were quantified from serum (29 biomarkers) or red blood cell samples (glycated haemoglobin; HbA1c) using 24 analysis methods across six analytical platforms from five manufacturers (AU5800, Beckman Coulter; AU5800, Randox; LIASON XL, DiaSorin Ltd.; VARIANT II Turbo, Bio-Rad; and ADVIA 1800, Siemens). Missing data arising due to biomarker concentrations being above or below limits of reportability or detection were replaced with the largest or smallest non-missing value of that biomarker respectively with a small offset of 0.0001 units. Further details on the 30 biomarkers can be found in **Table S3**.

#### Genotyping, imputation, and polygenic risk scores

UK Biobank participants were genotyped on UK BiLEVE arrays and UK Biobank Axiom arrays and imputed to the 1000 genomes, UK10K, and Haplotype Reference Consortium panels^60^ using human genome build GRCh37^27^. Here, we converted the data to PLINK2 probabilistic dosages^61^ for analyses.

PRSs for CHD and ischaemic stroke were obtained from the polygenic score (PGS) catalog^31^ (accessions PGS000018 and PGS000039 respectively) and computed in UK Biobank participants using the PLINK2 software^61^ (PLINK v2.00a3LM AVX2 Intel [2 Mar 2021]) linear scoring function applied to the probabilistic allele dosages. These PRSs were chosen for this study as they are the most predictive PRSs for CHD and ischaemic stroke that do not include GWAS summary statistics derived from UK Biobank participants in their model development.

#### SCORE2

Sex-specific per-participant values for SCORE2 (linear predictors) were computed from age, smoking status, SBP, total cholesterol, and HDL cholesterol using formulae published by the SCORE2 working group and ESC Cardiovascular risk collaboration^4^.

Age and sex at baseline assessment were obtained from UK Biobank fields #21003 and #31 respectively. Systolic blood pressure (SBP) was measured using either an automated digital device (OMRON) (field: #4080) and/or by manual sphygmomanometer (field: #93). In both cases, two measurements were taken several moments apart, and the average was taken to obtain a single representative measure of SBP. Smoking status was defined as “current” or “other” based on self-reported current smoking (field #20116). Missing data (“don’t know”, “prefer not to answer”) were set to “other”. The total and HDL cholesterol measurements used were those obtained by the clinical biochemistry assays as described above (see **Table S3** for UK Biobank field IDs).

Each risk factor was transformed as described in Supplementary Methods Table 2 of the SCORE2 publication^4^:

$$age=\left( age-60 \right)/5$$

$$smoking= \left\{ \begin{matrix} 1 if "current" \\ 0 if \text{"other" } \end{matrix} \right.$$

$$SBP=\left( SBP-120 \right)/{20}$$

$$tchol=\left( Total cholesterol-6 \right)/1$$

$$HDL=\left( HDL cholesterol-1.3 \right)/{0.5}$$

Then each transformed risk factor and risk factor × age- interaction was multiplied by the log hazard ratio obtained in the SCORE2 sensitivity analysis excluding UK Biobank participants from the log hazard ratio estimation (obtained from Supplementary Table 8 of the SCORE2 publication^4^ under the “Excluding UK Biobank” heading):

$${SCORE2}_{male}=\log\left( 1.50 \right) age+ \log\left( 1.77 \right)smoking+\log\left( 1.33 \right)SBP+\log\left( 1.13 \right)tchol+\log\left( 0.80 \right)HDL+\log\left( 0.92 \right)\left( age\times smoking \right)+\log\left( 0.98 \right)\left( age\times SBP \right)+\log\left( 0.98 \right)\left( age\times tchol \right)+\log\left( 1.04 \right)\left( age\times HDL \right)$$

$${SCORE2}_{female}=\log\left( 1.64 \right) age+ \log\left( 2.09 \right)smoking+\log\left( 1.39 \right)SBP+\log\left( 1.11 \right)tchol+\log\left( 0.81 \right)HDL+\log\left( 0.89 \right)\left( age\times smoking \right)+\log\left( 0.97 \right)\left( age\times SBP \right)+\log\left( 0.98 \right)\left( age\times tchol \right)+\log\left( 1.06 \right)\left( age\times HDL \right)$$

These log hazard ratios were used to prevent overestimation of the efficacy of SCORE2 for 10-year CVD risk prediction in this study (**Figure S9**), which could result in underestimation of any potential improvements in risk discrimination from addition of biomarkers or PRS. Results throughout were similar when using the main SCORE2 coefficients that included UK Biobank participants in their derivation (obtained from Supplementary Methods Table of the SCORE2 publication^4^), with modest attenuation as expected (data not shown).

The sex-stratified C-index for SCORE2 in the was computed directly from this SCORE2 linear predictor using the concordance function in the survival R package version 3.3-1. The standard error was computed using the infinitesimal jackknife method by the concordance function. The 95% confidence interval was computed assuming gaussian standard errors; by multiplying the standard error by the 2.5% and 97.5% percentiles of the normal distribution and adding the result to the absolute C-index.

#### QRISK3

Sex-specific per-participant linear predictor values for QRISK3 were computed using the algorithm published by ClinRisk Ltd. at https://qrisk.org/src.php, excluding the final step that converted the linear predictors to an absolute risk, which was applied separately as described in a later section of this **Supplementary Methods**.

Risk factors contributing to QRISK3 were: age, ethnicity, Townsend deprivation index, smoking status, SBP, standard deviation of SBP, height, weight, the ratio of total to HDL cholesterol, family history of myocardial infarction before the age of 60, atrial fibrillation, chronic kidney disease, erectile dysfunction, severe mental illness, migraines, rheumatoid arthritis, systemic lupus erythematosus, type 1 diabetes, type 2 diabetes, atypical antipsychotic medications, blood pressure treatment, and systemic corticosteroid usage^62^.

Age, sex, SBP, HDL and total cholesterol were extracted and processed as described above. Townsend deprivation index, height, and weight were extracted from UK Biobank fields #22189, #50, and #21002 respectively.

Self-reported ethnicity at baseline was extracted from UK Biobank field #21000 and recoded to match the ethnicity categories expected by the QRISK3 algorithm^62^. Participants self-reporting their ethnicity as “White” (code 1), “British” (code 1001), “Irish” (code 1002), “Any other White background” (code 1003), “Prefer not to answer”, had missing data in the ethnicity field were coded in the QRISK3 ethnicity category “White or not stated”. Participants self-reporting “Indian” (code 3001), “Pakistani” (code 3002), “Bangladeshi” (code 3003), “Black Caribbean” (code 4001), “Black African” (code 4002), or “Chinese” (code 5) were kept as-is as these categories matched QRISK3 ethnicity categories. Participants self-reporting “Asian or Asian British” (code 3) or “Any other Asian background” (code 3004) were coded in the QRISK3 ethnicity category “Other Asian”. All other self-reported ethnicities were coded in the QRISK3 category “Other ethnic group”.

Smoking status (non-smoker, ex-smoker, light smoker, moderate smoker, or heavy smoker) was defined based on both self-reported current smoking (field #20116) along with the number of cigarettes smoked per day for current smokers (field #3456). Participants with answering “don’t know” or “prefer not to answer” to the smoking status field (field #20116) were treated as non-smokers. Current smokers who reported smoking less than 10 cigarettes per day were coded as light smokers. Current smokers who reported smoking between 10 and 20 cigarettes per day were coded as moderate smokers. Current smokers who reported smoking 20 or more cigarettes per day were coded as heavy smokers. Current smokers with missing data in the number of cigarettes smoked per day field (field #3456) were treated as moderate smokers.

The standard deviation of SBP was calculated as the standard deviation of the two automated (field #4080) or manual (field #93) SBP measures depending on the data availability for the participant. If both automated and manual measures were taken, the mean of the automated and manual standard deviations was taken. If both the automated and manual measurements had only one measurement respectively, the standard deviation of SBP was calculated as the standard deviation of the manual and automated measurements.

Family history of myocardial infarction before the age of 60 in first degree relatives was treated as false for all participants, as the UK Biobank family history touchscreen questionnaire (field ID category #100034) did not include age of onset. Type 1 diabetes and type 2 diabetes status was also set to false, as participants with any history of diabetes did not meet the inclusion criteria for the study (see above).

History of atrial fibrillation was defined based on self-reported atrial fibrillation or atrial flutter during the interview with the trained nurse at baseline assessment (field #20002), or history of hospitalisation with atrial fibrillation or flutter (ICD-10 code I48, or ICD-9 codes 427.31 or 427.32).

Chronic kidney disease (CKD) status was defined following the criteria outlined in the QRISK3 publication^62^ to include end-stage renal disease (algorithmically defined outcome, field #42026), CKD stage 3 or higher (ICD­‑10 codes N18.3 or N18.4, or ICD-9 codes 585.3–585.6), nephrotic syndrome (ICD-10 code N04 or ICD-9 code 581), chronic glomerulonephritis (ICD-10 code N03 or ICD-9 code 582), chronic pyelonephritis (ICD-10 code N11 or ICD-9 code 590.0), dependence on renal dialysis (ICD-10 code Z99.2 or ICD-9 code V45.11), or history of kidney transplant (ICD-10 code Z94.0 or ICD-9 code V42.0). Note that the initial study criteria excluded participants with end-stage renal disease (see above). In total 107 participants were identified by this expanded CKD definition, who were retained in the study cohort.

History of severe mental illness included schizophrenia, bipolar disorder, and moderate/severe depression, which were identified based on a combination of self-reported mental health history in interview with trained nurse (field #20002), curated mental health status derived from touchscreen questionnaires returned by UK Biobank application 7155 (fields #20122, #20124, #20125, and #20126)^63^, and hospitalisation records (ICD‑10 codes F20, F31, or F32.1–F32.3, or ICD-9 codes 295, 296.0, 296.22–296.24, 296.4–296.7, or 298.0).

History of migraine was identified based on self-report during the interview with the trained nurse at baseline assessment (field #20002) and hospitalisation records (ICD-10 codes G43 or G44.0, or ICD-9 codes 339.0 or 346).

Rheumatoid arthritis was identified based on self-report during the interview with the trained nurse at baseline assessment (field #20002) and hospitalisation records (ICD-10 codes M05 or M06, or ICD-9 code 714).

Systemic lupus erythematosus was identified based on self-report during the interview with the trained nurse at baseline assessment (field #20002) and hospitalisation records (ICD-10 code M32, or ICD-9 code 710.0).

Erectile dysfunction status was identified based on self-reported erectile dysfunction during the interview with the trained nurse at baseline assessment (field #20002) and/or had been prescribed any of six medications to treat erectile dysfunction (**Table S19**) identified by cross-referencing the list of medications present in UK Biobank (field #2003) with the list of erectile dysfunction medications listed in the QRISK3 publication^62^.

Blood pressure lowering medication status was identified based on self-report in the touchscreen questionnaires (field #6177 for males, field #6153 for females) and/or had been prescribed any of 48 antihypertensive medications (**Table S19**) identified by cross-referencing the list of medications present in UK Biobank (field #20003) with the British National Formulary^59^ chapter 2.5: Hypertension and heart failure drugs with the restriction that the drug indications must include treatment of hypertension.

Systematic corticosteroid medication usage was identified where the participant had been prescribed any of nine medications (**Table S19**) identified by cross-referencing the list of medications present in UK Biobank (field #20003) against the British National Formulary^59^ chapter 6.3.2 Glucocorticoid therapy drugs as well as the list of corticosteroids in the QRISK3 publication^62^.

Atypical antipsychotic medication usage was identified where the participant had been prescribed any of eight medications (**Table S19**) identified by cross-referencing the list of medications present in UK Biobank (field #20003) with the list of atypical antipsychotic medications listed in the QRISK3 publication^62^.

Medical history and medication risk factors were set to false where data were missing or the participant answered “do not know” or “prefer not to answer” to the relevant touchscreen or verbal interview questions.

#### Incremental value in 10-year CVD risk prediction for individual biomarkers

Incremental improvement in 10-year CVD risk prediction for individual biomarkers beyond SCORE2 alone was assessed using differences in C-index from SCORE2 alone (ΔC-index) (**Figure 2**, **Table S4**). Incremental improvement in 10-year CVD risk was assessed for the 249 NMR biomarkers (**Table S2**) and 28 of the 30 clinical biochemistry assay biomarkers (**Table S3**): clinical biochemistry assays for HDL cholesterol and total cholesterol were not assessed here as they were used to compute SCORE2 linear predictor (see above).

In the discovery cohort (n=168,517; 5,096 CVD cases), for each biomarker we fit a sex-stratified Cox proportional hazards regression for 10-year CVD risk with the biomarker as an independent variable and SCORE2 as an offset term (**Figure 2B**, **Table S4**). SCORE2 was treated as an offset term, rather than an independent variable, as we sought to develop scores that added biomarkers to the existing SCORE2 weights. Cox proportional hazards regressions were fit using the coxph function in the survival R package version 3.3-1. The hazard ratio, absolute C-index, and its standard error were obtained directly from the returned result for each SCORE2 + biomarker model (**Table S4**). Hazard ratios were estimated per standard deviation increase in the respective biomarker.

In the replication cohort (n=128,946; 3,823 CVD cases; **Table S4**) and the pooled analysis (n=297,463; 8,919 CVD cases; **Figure 2A**), for each biomarker we predicted the model fit by (1) standardising the biomarker to have mean 0 and standard deviation 1, (2) multiplying by the log hazard ratio estimated in the discovery cohort, then (3) adding the result to the SCORE2 linear predictor. The absolute C-index and its standard error were computed on the combined linear predictor using the concordance function. Means and standard deviations for each biomarker were similar in the discovery and replication cohorts (**Table S4**).

The ΔC-index was subsequently computed for each SCORE2 + biomarker model by subtracting the sex-stratified C-index for the SCORE2 linear predictor as described above. For each biomarker, the standard error of the ΔC-index was calculated accounting for the covariance between the C-index standard errors:

$$\sigma_{\Delta\text{C-index}}=\sqrt{\sigma_{\text{SCORE2 C-index}}^{2}+\sigma_{\text{SCORE2+biomarker C-index}}^{2}-2\sigma_{\text{SCORE2 C-index},\text{SCORE2+biomarker C-index}}}$$

95% confidence intervals for the ΔC-index were calculated by multiplying the standard error by the 2.5% and 97.5% percentiles of the normal distribution. Two-sided p-values were computed after converting the ΔC-index into a Z-score by dividing the ΔC-index by its standard error. P-values were corrected for multiple testing across the 277 tested biomarkers using Benjamini-Hochberg false-discovery rate (FDR) correction.

In addition to sex-stratified analysis, incremental improvements in C-index beyond SCORE2 alone were similarly assessed in sex-specific analysis (**Table S5**, **Figure S2**); performed separately in the 127,269 male participants (5,633 incident CVD cases) and in the 170,194 female participants (3,286 incident CVD cases). Analyses were performed as described above without the sex-stratification term. Likewise, analyses were performed as described above for sensitivity analyses to the narrow incident CVD definition and when using QRISK3 in place of SCORE2 (**Table S5**, **Figure S2**).

#### NMR biomarker score training

Sex-specific NMR biomarker scores for 10-year risk of CHD (4,054 cases; **Table S20A**) and 10-year risk of ischaemic stroke (1,280 cases; **Table S20B**) were trained and tested in the 168,517 participants in the discovery cohort, then later combined (see next section) for 10-year CVD risk prediction (**Figure S10**). NMR biomarker scores were trained for CHD and stroke separately and later combined, as we previously found that combining PRSs for CHD and stroke led to improved prediction of 10-year CVD risk over a single PRS trained for CVD, due to its heterogeneity^12^. NMR scores were also trained in males and females separately to capture sex-specific differences in their concentrations^30^ and well-known differences in baseline survival between males and females^3^. NMR scores were trained using elastic-net penalised Cox proportional hazards regression^64,65^ in nested cross validation using the 106 non-derived NMR biomarkers as candidate predictors (**Figure S6**). Biomarkers were classified as non-derived where they could not be computed by summing or dividing two or more other biomarkers^30^ (**Table S2**). The per-biomarker weights for computing the consensus optimal NMR scores after model training are given in **Table S6**.

Incident CHD was defined as fatal or non-fatal myocardial infarction (ICD-10 codes I21–I24 or I25.2) or by presence of major coronary surgery (ICD-10 codes Z95.1 or OPSC-4 codes K40–K46, K49, K50.1, K75) within 10-years of follow-up. Incident ischaemic stroke was determined using the respective UK Biobank algorithmically defined outcome (field #42008) within 10-years of follow-up. Follow-up time was defined for each endpoint separately; *i.e.,* a non-fatal ischaemic stroke event prior to CHD was not counted as a competing risk when defining follow-up for incident CHD and *vice versa*. Case numbers and follow-up characteristics are reported in **Table S20A** for CHD and **Table S20B** for ischaemic stroke.

Prior to NMR biomarker score training, missing data in the 106 NMR biomarkers were imputed as glmnet could not handle missing data. For model training purposes, missing data were imputed a single time in the 168,517 participants in the discovery cohort using the impute R package version 1.70.0 with the K-nearest neighbours algorithm^66^. K was set to 20 based on the number of principal components that cumulatively explained >95% of the variation in the 106 non-derived biomarkers. Prior to imputation, >92% of participants had no missing NMR biomarker concentrations, >6% had only one biomarker missing, and the remaining <2% had 2–5 of 106 biomarkers missing. In total 0.1% of biomarker concentrations were imputed. Per-biomarker missingness rates are given in **Table S2**. Biomarker concentrations were standardised in males and females separately so that sex-specific coefficients fit by glmnet were comparable across biomarkers.

When training NMR scores using elastic-net penalised Cox proportional hazards, a nested cross-validation procedure was used, comprising a 5-fold outer layer and 10-fold inner layer (**Figure S10**). For each iteration at the outer layer, NMR biomarker scores were trained in 4/5^ths^ of the data, then predicted in the withheld 1/5^th^ of the data (test fold) (**Figure S10A**). Model training in each iteration was performed with elastic-net penalised Cox proportional hazards regression using the glmnet R package version 4.1-6 (**Figure S10A**). Each NMR biomarker score was trained with the 106 non-derived NMR biomarkers with SCORE2 as an offset term. Within each iteration, 10-fold cross-validation (inner-layer) for hyperparameter tuning of the elasticnet model (**Figure S10B**). Random allocation of participants to test folds was performed using the caret R package version 6.0-92 to balance the number of males and females, and incident cases within males and females, across test folds. Test folds at the inner cross-validation layer were balanced by incident CHD or ischaemic stroke cases respectively when training NMR scores for the respective endpoint. Test-folds at the outer cross-validation layer were balanced by incident CVD cases as the outer cross-validation layer was also used for the training procedure used to combine NMR scores for CHD and stroke described in the next section.

For each of the five training iterations, the optimal elastic-net fit was determined with a grid search of α and λ where the range of α was [0 (ridge regression), 0.1, 0.25, 0.5, 0.75, 0.9, and 1 (lasso regression)] and a sequence of 100 λ values was automatically determined by the glmnet function for each α (**Figure S11A**). β coefficients estimated for each biomarker for the optimal fit in each training data are shown in **Figure S11C**. The proportion of variance in the NMR score explained by each biomarker was calculated as the ratio of its β^2^ to the sum of β^2^ across all biomarkers. To obtain a single representative score (*i.e.,* for computing NMR scores in new samples), coefficients were averaged across the five training iterations (**Figures S11C**, **Table S6**). Proportions of variance explained in the NMR score by each biomarker were also averaged across the five training iterations (**Table S6**).

All four consensus optimal NMR scores included all 106 non-derived biomarkers (**Figure S11C**, **Table S6**). The biomarker with the strongest contribution to the CHD NMR scores was GlycA, which explained 12.8% of the variance in the CHD NMR score in males, and 11.4% of the variance in the CHD NMR score in females (**Table S6**). The biomarker with the strongest contribution to the ischaemic stroke NMR scores was albumin, which explained 15.7% of the variance in the ischaemic stroke NMR score in males, and 27.1% of the variance in the ischaemic stroke NMR score in females (**Table S6**).

To avoid overestimation of prediction performance in downstream analyses in the discovery cohort, for each NMR score we used the aggregate of their predicted values across cross-validation test partitions (**Figure S10A**). Sex-specific pairwise correlations between SCORE2, the NMR scores, and PRSs are shown in **Figure S12**.

#### Incremental value in 10-year CVD risk prediction for biomarker combinations and PRSs

NMR biomarker scores and FDR-significant clinical biochemistry biomarkers were assessed for incremental improvement in 10-year CVD risk prediction over SCORE2 in combination with PRSs. In total, we compared five models to SCORE2: (1) SCORE2 + clinical biomarkers, (2) SCORE2 + NMR scores, (3) SCORE2 + PRSs, (4) SCORE2 + clinical biomarkers + PRSs, and (5) SCORE2 + NMR scores + PRSs. For each model, we fit multivariable Cox proportional hazards models for 10-year CVD with SCORE2 as an offset term in the discovery cohort after standardising each clinical biochemistry biomarker, NMR score, and PRS. Multivariable models were fit in males and females separately in order to estimate sex-specific coefficients for biomarkers/NMR scores/PRSs that could be added to the sex-specific SCORE2 models (see above) and later combined with sex-specific formulae for converting SCORE2 linear predictors into absolute risks (see below). Sex-specific coefficients for each biomarker/NMR score/PRS in each model that can be used when predicting model fits in new samples are shown in **Figure 2** and detailed in **Table S7**. Associations between clinical biomarkers, NMR scores, and PRSs in the context of individual SCORE2 and QRISK3 risk factors are shown in **Figure S13** and **Table S21**.

In the replication cohort (**Table S8**) and pooled analysis (**Figure 4A**) predicted model fits were obtained by multiplying each standardised biomarker/NMR score/PRS by their model-specific coefficients and adding the result to the SCORE2 linear predictor. Means and standard deviations for each biomarker/NMR score/PRS were similar in the discovery and replication cohorts (**Table S8**). Sex-stratified C-indices, ΔC-indices, standard errors, 95% confidence intervals, and p-values were computed from resulting linear predictors as described above.

In addition to sex-stratified analysis, incremental improvements in C-index beyond SCORE2 alone were similarly assessed in sex-specific analysis (**Table S9**, **Figure S3**) as well as in sensitivity analyses to the narrow incident CVD definition, and when using QRISK3 in place of SCORE2 (**Table S9**, **Figure S3**).

#### Computation of absolute risks

Linear predictors for SCORE2, SCORE2 + NMR scores, SCORE2 + clinical biomarkers, SCORE2 + PRSs, SCORE2 + NMR scores + PRSs, and SCORE2 + clinical biomarkers + PRSs were converted into predictions of absolute 10-year CVD risk using formulae developed by the SCORE2 working group and ESC cardiovascular risk consortium^4^:

First, uncalibrated absolute risks were calculated using sex-specific estimates of baseline survival, which they calculated as the median baseline survival across 44 cohorts (including UK Biobank)^4^:

$${Uncalibrated 10year risk}_{Males}=1-{0.9605}^{\exp\left( linear predictor \right)}$$

$${Uncalibrated 10year risk}_{Females}=1-{0.9776}^{\exp\left( linear predictor \right)}$$

Second, these were converted into absolute 10-year risks calibrated to the UK population using their scaling factors for the low-risk European region (which included the UK) applied in their risk recalibration formula^4^:

$${Calibrated 10year risk}_{Males}=1-exp\left( -exp\left( -0.5699+0.7476\times ln\left( -ln\left( 1-Uncalibrated 10year risk \right) \right) \right) \right)$$

$${Calibrated 10year risk}_{Females}=1-exp\left( -exp\left( -0.7380+0.7019\times ln\left( -ln\left( 1-Uncalibrated 10year risk \right) \right) \right) \right)$$

Distributions of absolute risk for each model are shown in **Figure S14** and were similar in both the discovery cohort, where models were fit, and in the replication cohort, where model fits were predicted.

Similarly, linear predictors for QRISK3, QRISK3 + NMR scores, QRISK3 + clinical biomarkers, QRISK3 + PRSs, QRISK3 + NMR scores + PRSs, and QRISK3 + clinical biomarkers + PRSs were converted into predictions of absolute 10-year CVD risk using the sex-specific estimates of baseline survival obtained by the QRISK3 authors using primary care data from England^62^ and published by ClinRisk Ltd. at https://qrisk.org/src.php:

$${Calibrated 10year risk}_{Males}=1-{0.9773}^{\exp\left( linear predictor \right)}$$

$${Calibrated 10year risk}_{Females}=1-{0.9889}^{\exp\left( linear predictor \right)}$$

Distributions of absolute risk for each model are shown in **Figure S15** and were similar in both the discovery cohort, where models were fit, and in the replication cohort, where model fits were predicted.

#### Incremental value in 10-year CVD risk stratification at risk thresholds used for clinical decision making

Incremental improvement in 10-year CVD risk stratification at risk thresholds used for clinical decision making was assessed by categorical NRI analysis^32,33^ (**Figure 4B**, **Table S10–S11**). When assessing incremental improvement over SCORE2, participants were stratified into low, medium, and high-risk groups as described in the **Methods**; based on their model-specific predicted absolute risk (see above) using age-specific risk thresholds recommended by the ESC 2021 guidelines for treatment consideration^3^. When assessing incremental improvement over QRISK3 in the sensitivity analyses (**Figure S4**, **Table S12**–**S13**), participants were stratified into a low and high-risk groups using a risk threshold of 10% recommended by the UK’s National Institute for Health and Care Excellence (NICE) 2023 guidelines for cardiovascular risk assessment and reduction^36^. Categorical net reclassification estimates were obtained using the nricens R package version 1.6. Standard errors were calculated in a bootstrap procedure with 1,000 bootstraps. 95% confidence intervals and two-sided P-values were computed from the bootstrap standard error using the first order normal approximation method.

#### Incremental value for CVD prevention for population-wide and targeted screening

Incremental benefits for primary CVD prevention if applied to the UK primary care population eligible for screening were assessed in population-wide and targeted screening strategies (**Figure 5**, **Table S15**) as outlined in the **Methods**. In both cases, study participants were stratified into low, medium, and high-risk groups based on their predicted absolute risks as described above. When assessing improvements in CVD prevention over QRISK3 in the targeted screening strategy (**Figure S8**, **Table S16**), the medium risk group was defined as people with >5% risk. Numbers of CVD cases and non-cases stratified into each risk group were standardised to the demographics and expected CVD incidents rates of the UK primary care population eligible for screening as outlined in the **Methods**. As expected^26^, the UK Biobank study cohort was older and healthier than the UK primary care population eligible for screening (**Figure S16**, **Table S22**).

A schematic showing a worked example of the standardisation procedure is shown in **Figure S17**. For each model and screening strategy, proportions of CVD cases and non-cases in each risk strata were calculated in males and females and five-year age groups. These proportions were then multiplied by numbers per 100,000 individuals in the general UK population aged 40–69 in the respective age- and sex- strata and the expected 10-year CVD incidence rates in each respective age- and sex- strata amongst CVD- and statin- free primary care patients (**Figure S18**, **Table S22**).

Sex-specific CVD incidence rates expected in each five-year age-group were obtained from Table A in the S2 Text in Sun *et al.* 2021^12^. These CVD incidence rates were obtained by Sun *et al.* from a random sample of 2.1 million people amongst 11.3 million CVD- and statin- free primary care patients 35–74 years of age who, between 2004–2017, attended any of 674 general practices opting into data linkage to the UK Clinical Practice Research Datalink (CPRD)^67^. The published per-1000-year CVD incidence rates in the five-years ahead (Table A in the S2 Text in Sun *et al.* 2021^12^) were converted into percentages of males and females expected to have an incident CVD event within 10 years as 1−exp(rate/1000) (**Figure S18**). These percentages were applied to numbers in each age- and sex- group in the general UK population obtained from the mid-2023 UK population estimates published by the Office for National Statistics^35^, then converted to numbers of CVD cases and non-cases per 100,000 (**Figure S18**).

The impact of statin initiation was modelled as preventing one in five incident CVD events based on evidence from multiple large clinical trials that have found that a 1 mmol/L reduction in LDL cholesterol with standard statin regimens reduces 5-year relative risk of major vascular events by 20% independent of patient health, absolute risk, and starting LDL cholesterol^34,68,69^.

Standard errors for N_high-risk_, CVD_high-risk_, CVD_prevented_, NNS, NNT (**Methods**) for each model and screening strategy and their Δ’s relative to SCORE2 alone were calculated in a bootstrap procedure with 1,000 bootstraps using the censboot function in the boot R package (version 1.3-28) with methods appropriate for right-censored data^70^. 95% confidence intervals and two-sided P-values were computed from the bootstrap standard error using the first order normal approximation method.

### Supplementary Figures

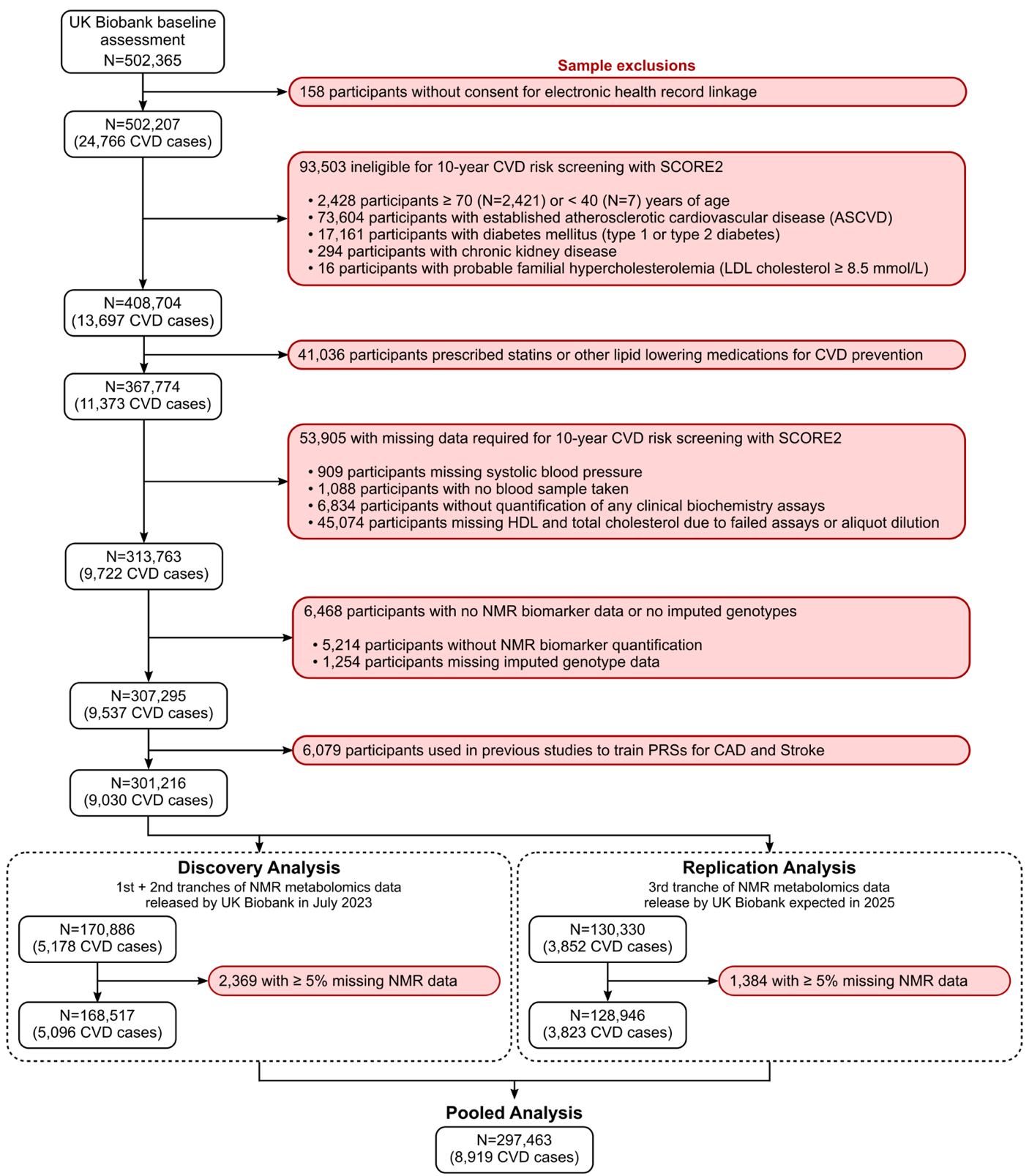

Figure S1: Sample inclusion and exclusion criteria and flowchart to arrive at study cohorts

Established atherosclerotic cardiovascular disease included prior history of heart disease, stroke, transient ischaemic attack, peripheral vascular disease, angina, and history of coronary surgery (**Table S1**). Presence and likelihood of familial hypercholesterolemia was assessed using the Dutch Lipids Clinical Network diagnostic criteria score (**Supplementary Methods**). CVD cases: cardiovascular disease events within 10 years of follow-up.

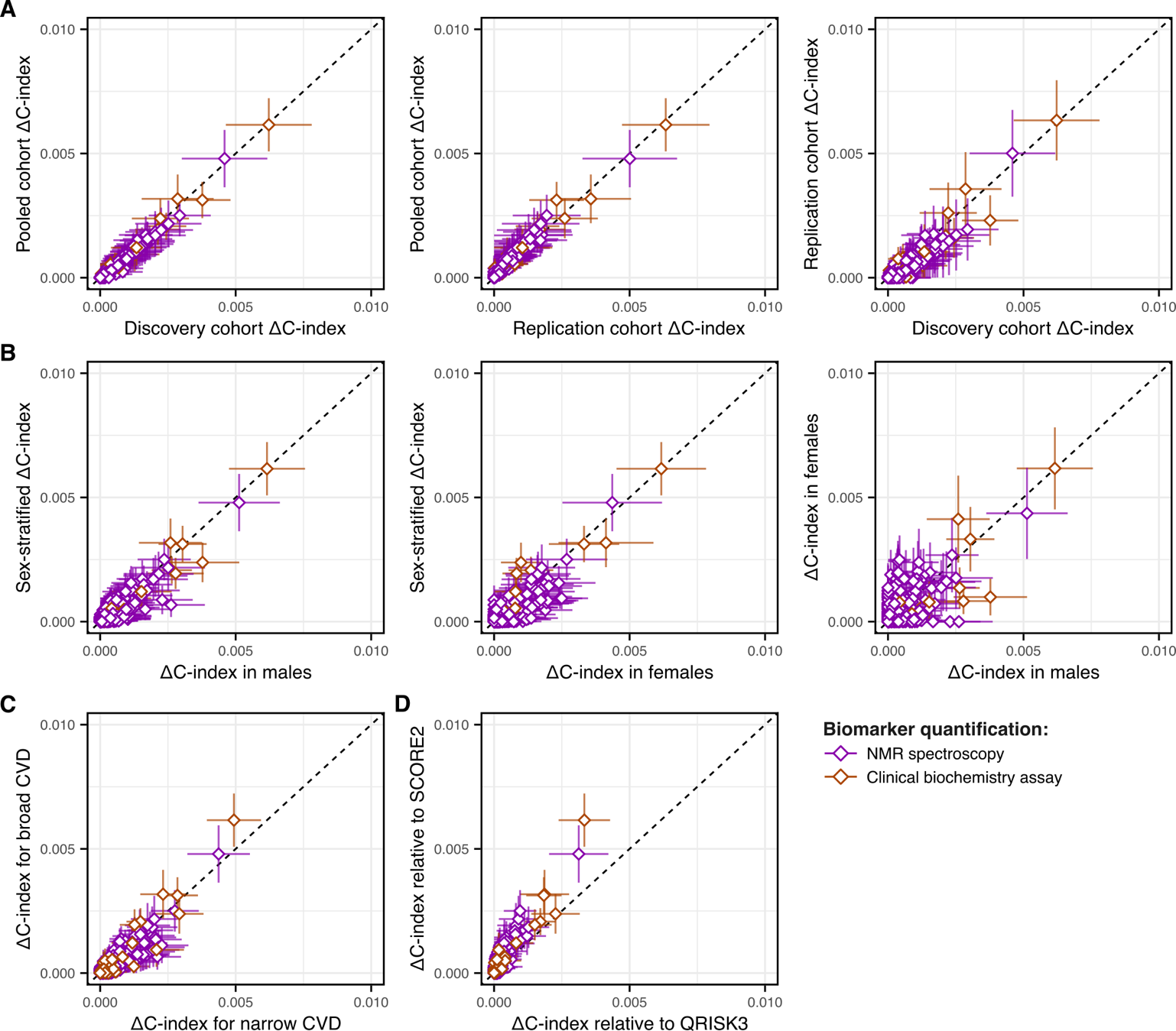

Figure S2: Sensitivity analysis of ΔC-index for individual biomarkers

**A)** Comparison of ΔC-index obtained for the 277 biomarkers across the discovery, replication, and pooled analysis cohorts. **B)** Comparison of ΔC-index obtained for the 277 biomarkers across sex-stratified and sex-specific analyses. **C)** Comparison of ΔC-index obtained for the 277 biomarkers when using the different CVD definitions. **D)** Comparison of ΔC-index obtained for the 277 biomarkers when using SCORE2 vs. QRISK3 as the conventional risk score. **A–D)** Horizontal and vertical bars show the 95% confidence intervals for the ΔC-index in the respective analyses. Further details on the sensitivity analyses are provided in the **Methods** and technical details on the computation of the ΔC-index in the **Supplementary Methods**. ΔC‑indices and related statistical information for all sensitivity analyses are reported in **Table S5**.

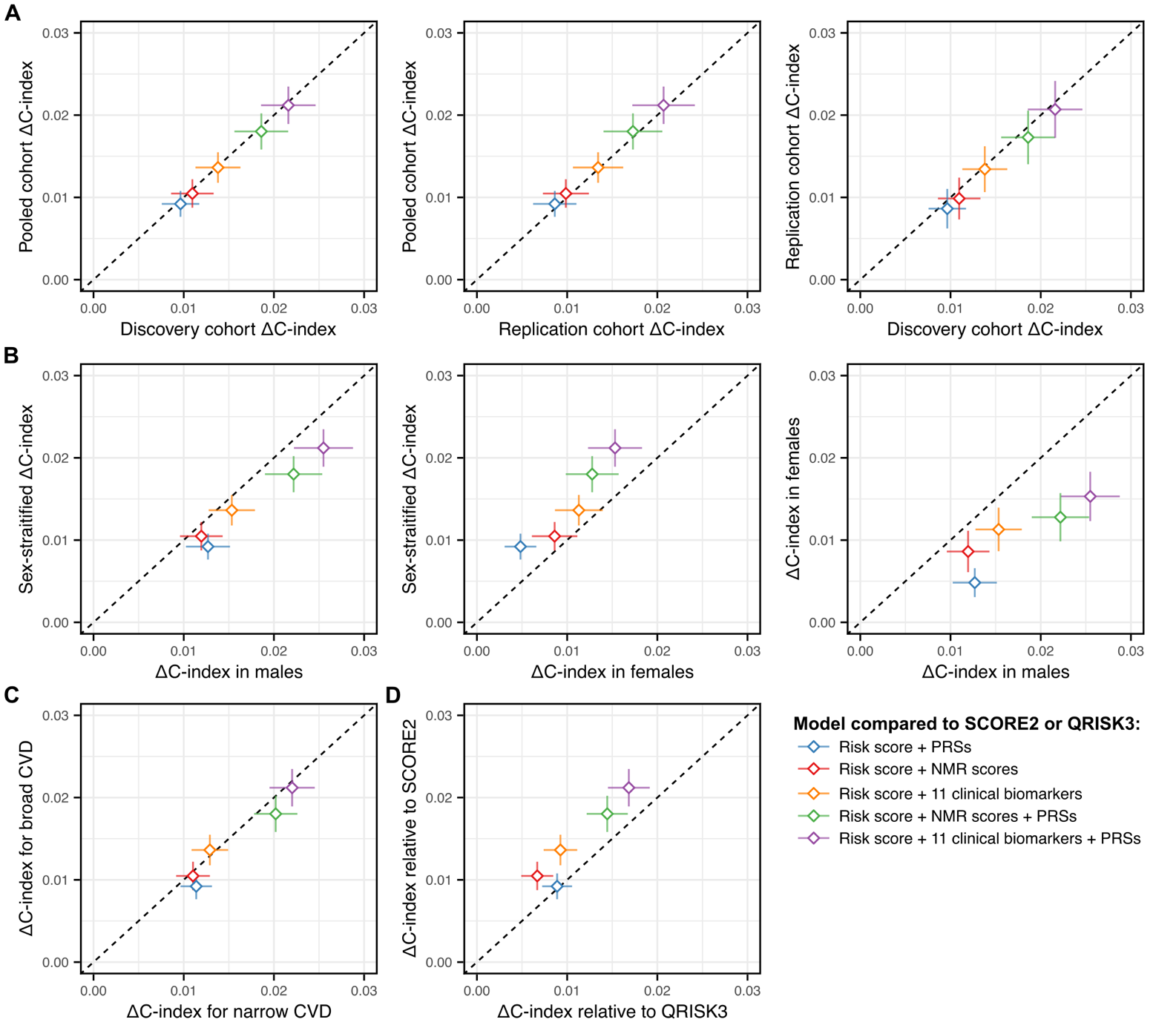

Figure S3: Sensitivity analysis of ΔC-index for NMR scores, combined biomarkers, and PRS

**A)** Comparison of ΔC-index across the discovery, replication, and pooled analysis cohorts for the five models adding NMR scores, clinical biomarkers, and/or PRSs to SCORE2. **B)** Comparison of ΔC-index across sex-stratified and sex-specific analyses for the five models adding NMR scores, clinical biomarkers, and/or PRSs to SCORE2. **C)** Comparison of ΔC-index when using the different CVD definitions for the five models adding NMR scores, clinical biomarkers, and/or PRSs to SCORE2. **D)** Comparison of ΔC-index when adding NMR scores, clinical biomarkers, and/or PRSs to SCORE2 vs adding NMR scores, clinical biomarkers, and/or PRSs to QRISK3. **A–D)** Horizontal and vertical bars show the 95% confidence intervals for the ΔC-index in the respective analyses. Further details on the sensitivity analyses are provided in the **Methods** and technical details on the computation of the ΔC-index in the **Supplementary Methods**. ΔC‑indices and related statistical information for all sensitivity analyses are reported in **Table S9**.

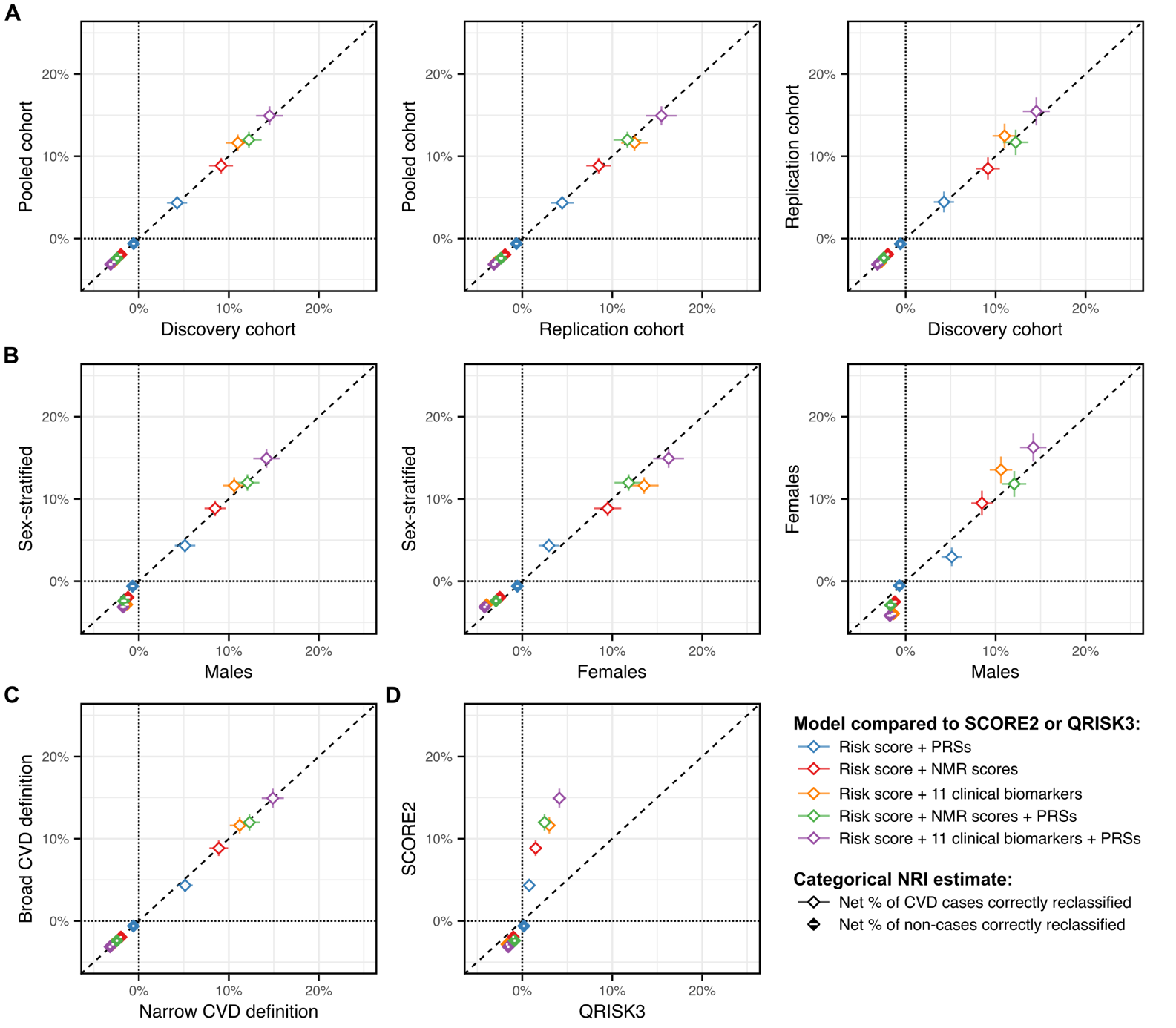

Figure S4: Sensitivity analysis of categorical NRI for NMR scores, combined biomarkers, and PRSs

**A)** Comparison of net % of CVD cases and non-cases correctly reclassified across the discovery, replication, and pooled analysis cohorts when comparing SCORE2 to the five models adding NMR scores, clinical biomarkers, and/or PRSs to SCORE2. **B)** Comparison of net % of CVD cases and non-cases correctly reclassified across sex-stratified and sex-specific analyses when comparing SCORE2 to the five models adding NMR scores, clinical biomarkers, and/or PRSs to SCORE2. **C)** Comparison of net % of CVD cases and non-cases correctly reclassified when using the different CVD definitions when comparing SCORE2 to the five models adding NMR scores, clinical biomarkers, and/or PRSs to SCORE2. **D)** Comparison of net % of CVD cases and non-cases correctly reclassified when adding NMR scores, clinical biomarkers, and/or PRSs to SCORE2 vs adding NMR scores, clinical biomarkers, and/or PRSs to QRISK3. **A–D)** Horizontal and vertical bars show the 95% confidence intervals for the net % correctly reclassified in the respective analyses. Further details on the sensitivity analyses are provided in the **Methods** and technical details on the categorical NRI analyses in the **Supplementary Methods**. Net % of CVD cases and non-cases correctly reclassified and related statistical information for all sensitivity analyses are reported in **Table S12** and numbers allocated to each risk category by each model are detailed in **Table S13**.

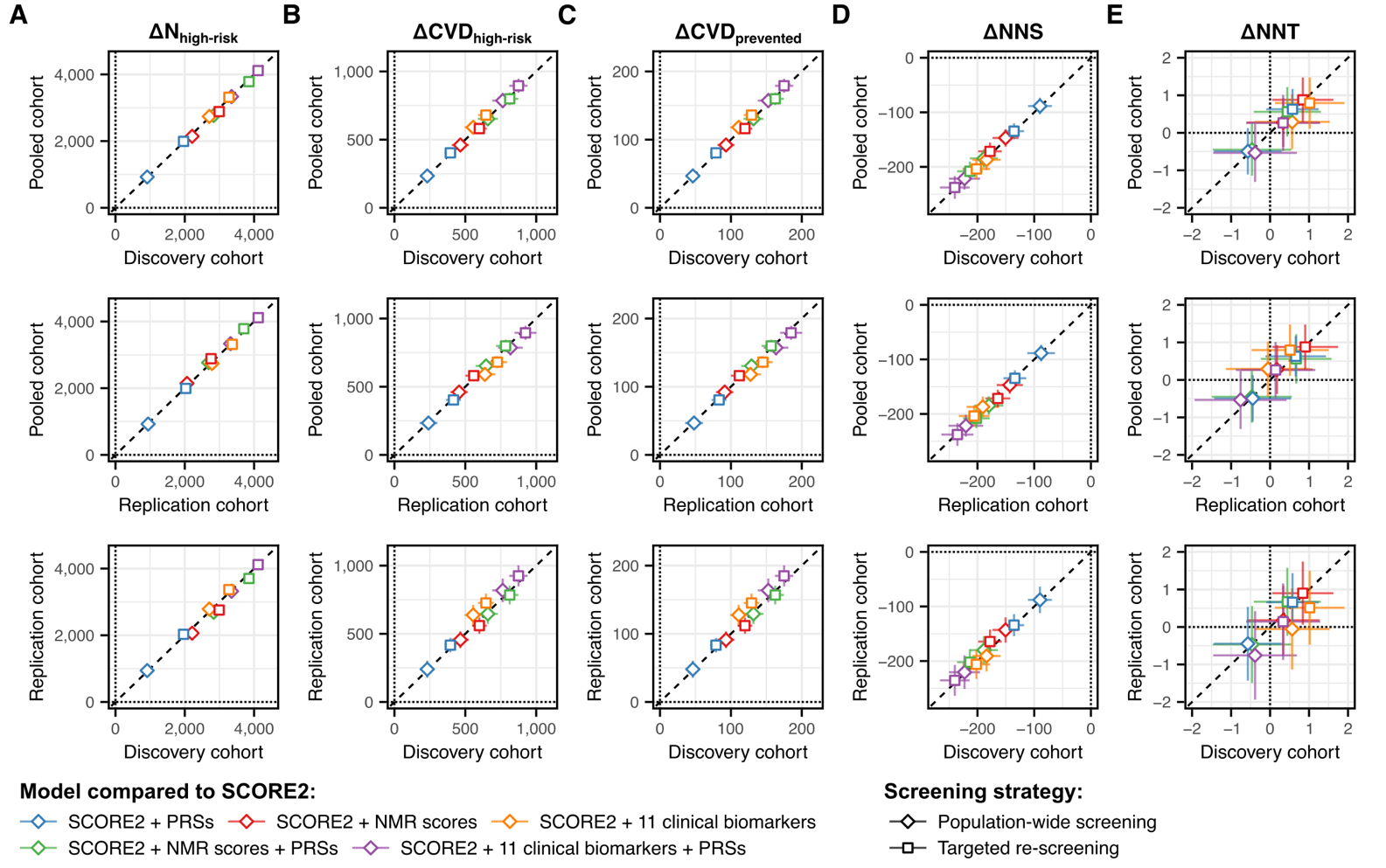

Figure S5: Sensitivity analysis of incremental value for CVD prevention to study cohort

Comparison of incremental value for primary prevention of CVD per 100,000 in the UK eligible for screening when deriving estimates from risk stratification in the discovery, replication, and pooled analysis cohorts. **A)** Comparison of the change in number classified as high risk (ΔN_high-risk_). **B)** Comparison of the change in number of incident CVD cases amongst the high-risk group (ΔCVD_prevented_). **C)** Comparison of the change in number of CVD events expected be prevented within 10 years by initiation of statins in the high-risk group (ΔCVD_prevented_). **D)** Comparison of the change in the number needed to screen to prevent one CVD event (ΔNNS). **E)** Comparison of the change in number the number of statins prescribed per CVD event prevented (ΔNNT). **A–E**) Horizontal and vertical bars show the 95% confidence intervals for each statistic. Further details on the sensitivity analyses are provided in the **Methods** and technical details in the **Supplementary Methods**. Point estimates and 95% confidence intervals are reported in **Table S16**.

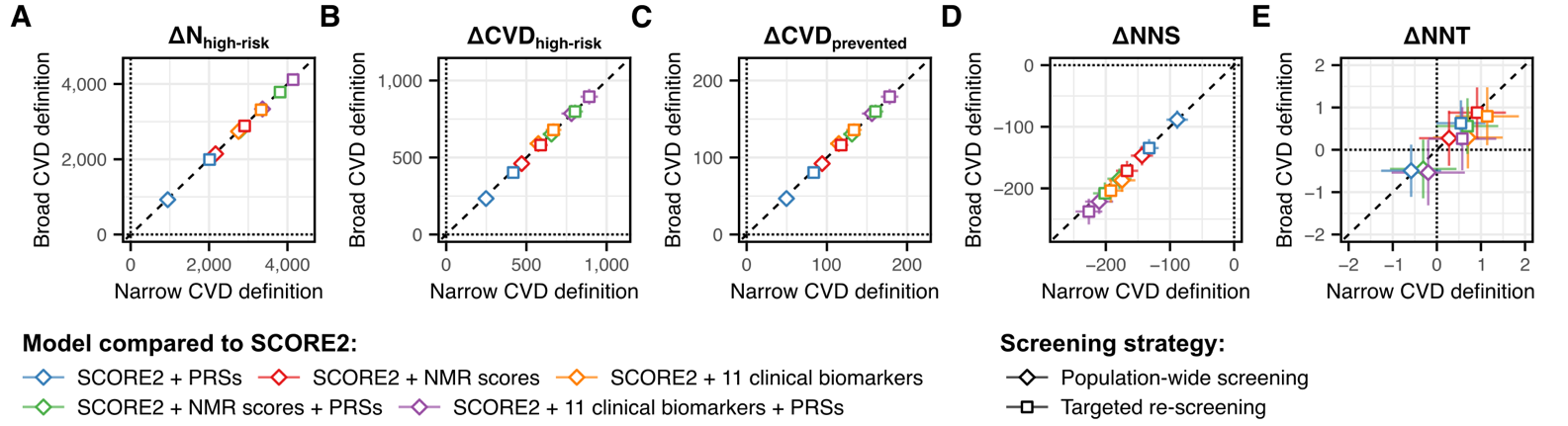

Figure S6: Sensitivity analysis of incremental value for CVD prevention to CVD definition

Comparison of incremental value for primary prevention of CVD per 100,000 in the UK eligible for screening when deriving estimates from risk stratification in the study cohort using different CVD definitions (**Methods**). **A)** Comparison of the change in number classified as high risk (ΔN_high-risk_). **B)** Comparison of the change in number of incident CVD cases amongst the high-risk group (ΔCVD_prevented_). **C)** Comparison of the change in number of CVD events expected be prevented within 10 years by initiation of statins in the high-risk group (ΔCVD_prevented_). **D)** Comparison of the change in the number needed to screen to prevent one CVD event (ΔNNS). **E)** Comparison of the change in number the number of statins prescribed per CVD event prevented (ΔNNT). **A–E**) Horizontal and vertical bars show the 95% confidence intervals for each statistic. Further details on the sensitivity analyses are provided in the **Supplementary Methods**. Point estimates and 95% confidence intervals are reported in **Table S16**.

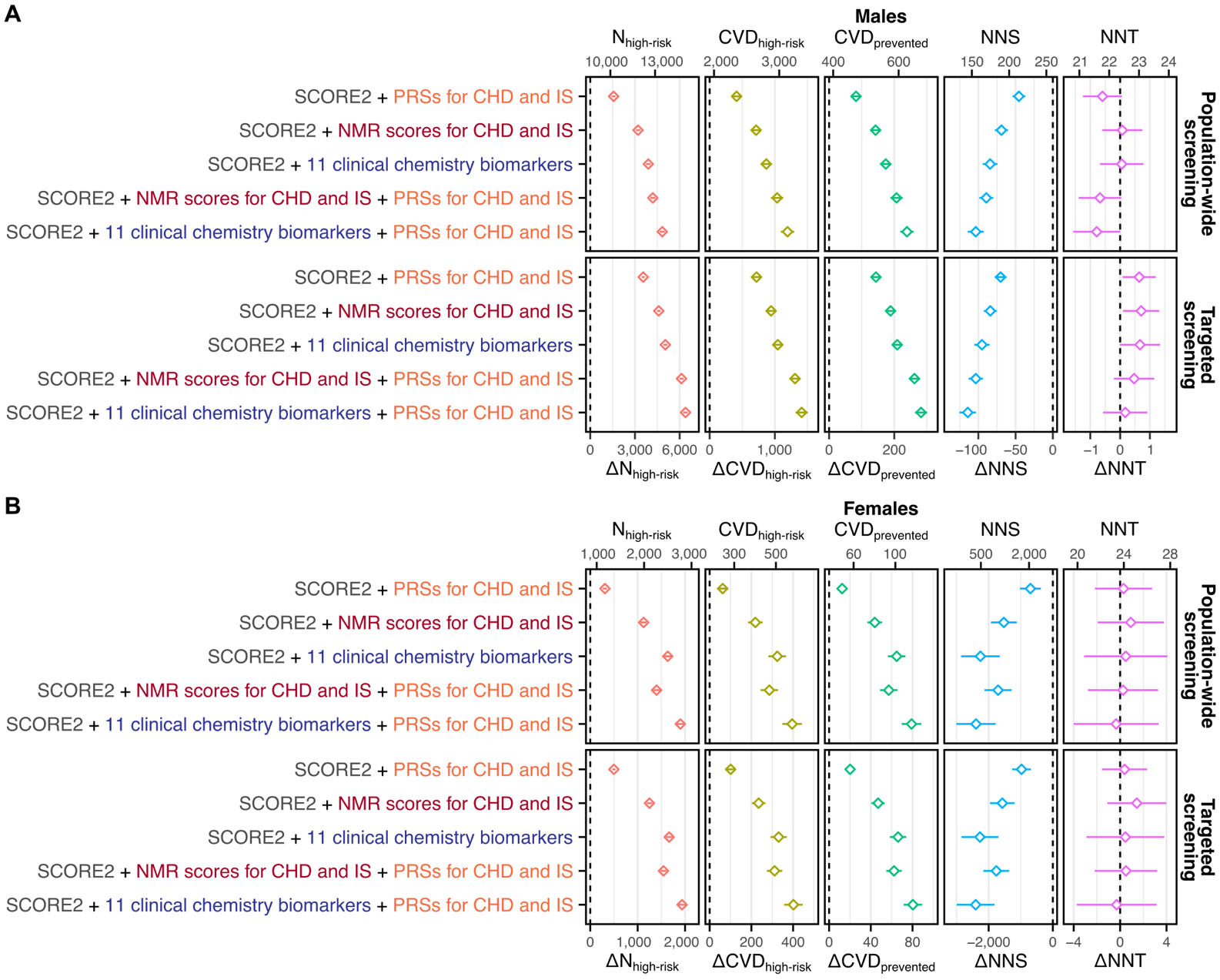

Figure S7: Incremental value for CVD prevention in males and females separately

**A)** Incremental value for primary prevention of CVD per 100,000 males (9,025 CVD cases) in the UK eligible for screening when adding NMR scores, clinical biomarkers, and/or PRSs to SCORE2 in population-wide or targeted screening. **B)** Incremental value for primary prevention of CVD per 100,000 females (4,447 CVD cases) in the UK eligible for screening when adding NMR scores, clinical biomarkers, and/or PRSs to SCORE2 in population-wide or targeted screening. **A–B**) 95% confidence intervals were estimated via a bootstrap sampling procedure with 1000 bootstraps (**Methods**). Further details on the sensitivity analyses are provided in the **Supplementary Methods**. Point estimates and 95% confidence intervals are detailed in **Table S16**.

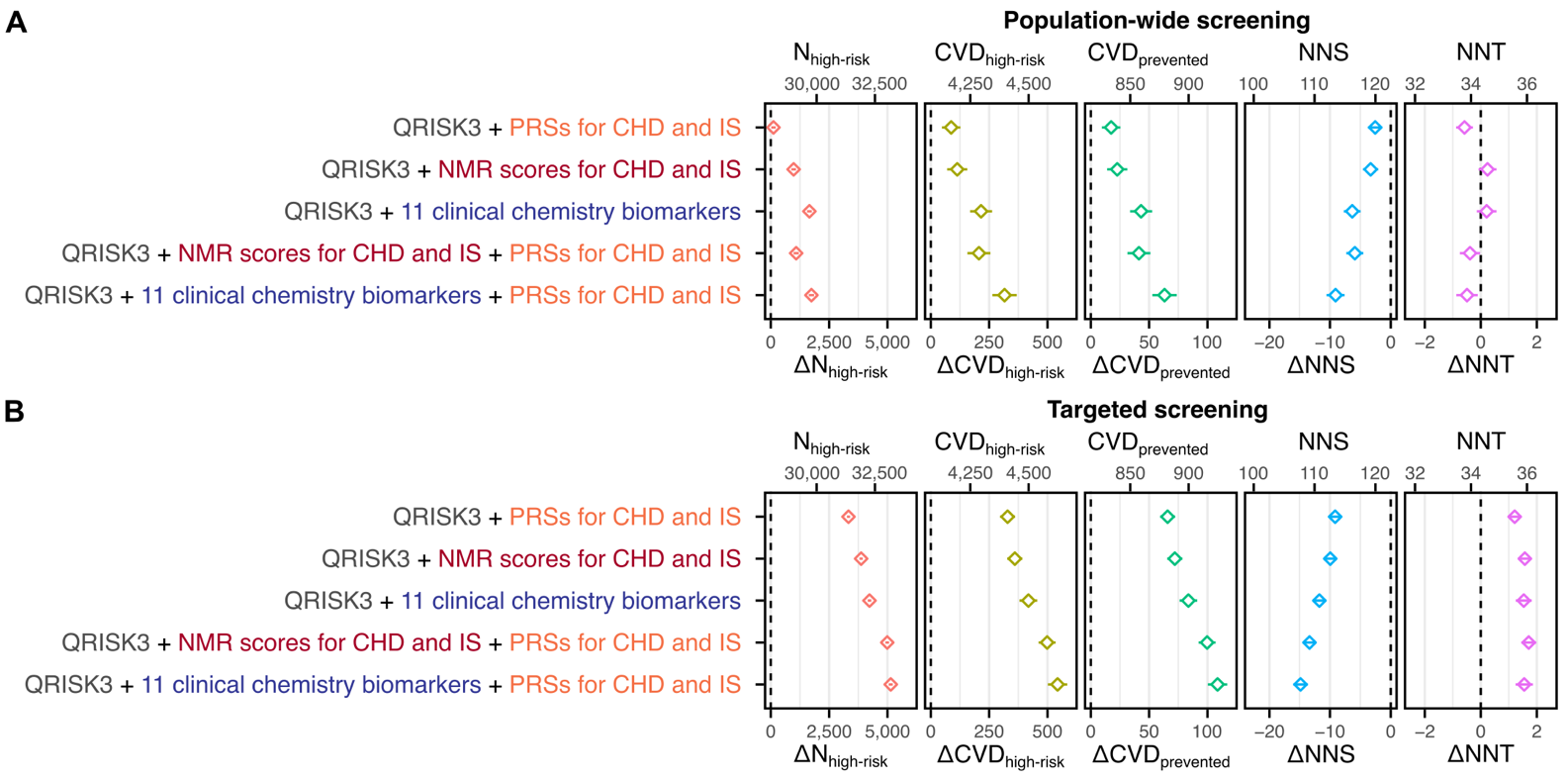

Figure S8: Incremental value for CVD prevention when using QRISK3

**A)** Incremental value for primary prevention of CVD per 100,000 in the UK eligible for screening when adding NMR scores, clinical biomarkers, and/or PRSs to QRISK3. **B)** Incremental value for primary prevention of CVD when using NMR scores, clinical biomarkers, and/or PRSs for targeted risk-reclassification in people classified as medium risk by QRISK3 alone. **A–B**) 95% confidence intervals were estimated via a bootstrap sampling procedure with 1000 bootstraps (**Methods**). Further details on the sensitivity analyses are provided in the **Supplementary Methods**. Point estimates and 95% confidence intervals are detailed in **Table S16**.

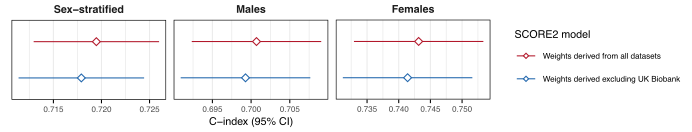

Figure S9: Prevention of overfitting of SCORE2 in UK Biobank participants.

Compares the C-index and its 95% confidence interval (95% CI) for 10-year CVD risk in the discovery cohort (n=168,517; 5,096 CVD cases) obtained using SCORE2 when computed from its published coefficients that were derived with (red) and without (blue) UK Biobank participants. For all analyses in the study, SCORE2 was derived using the per- risk factor weightings derived excluding UK Biobank (**Supplementary** **Methods**).

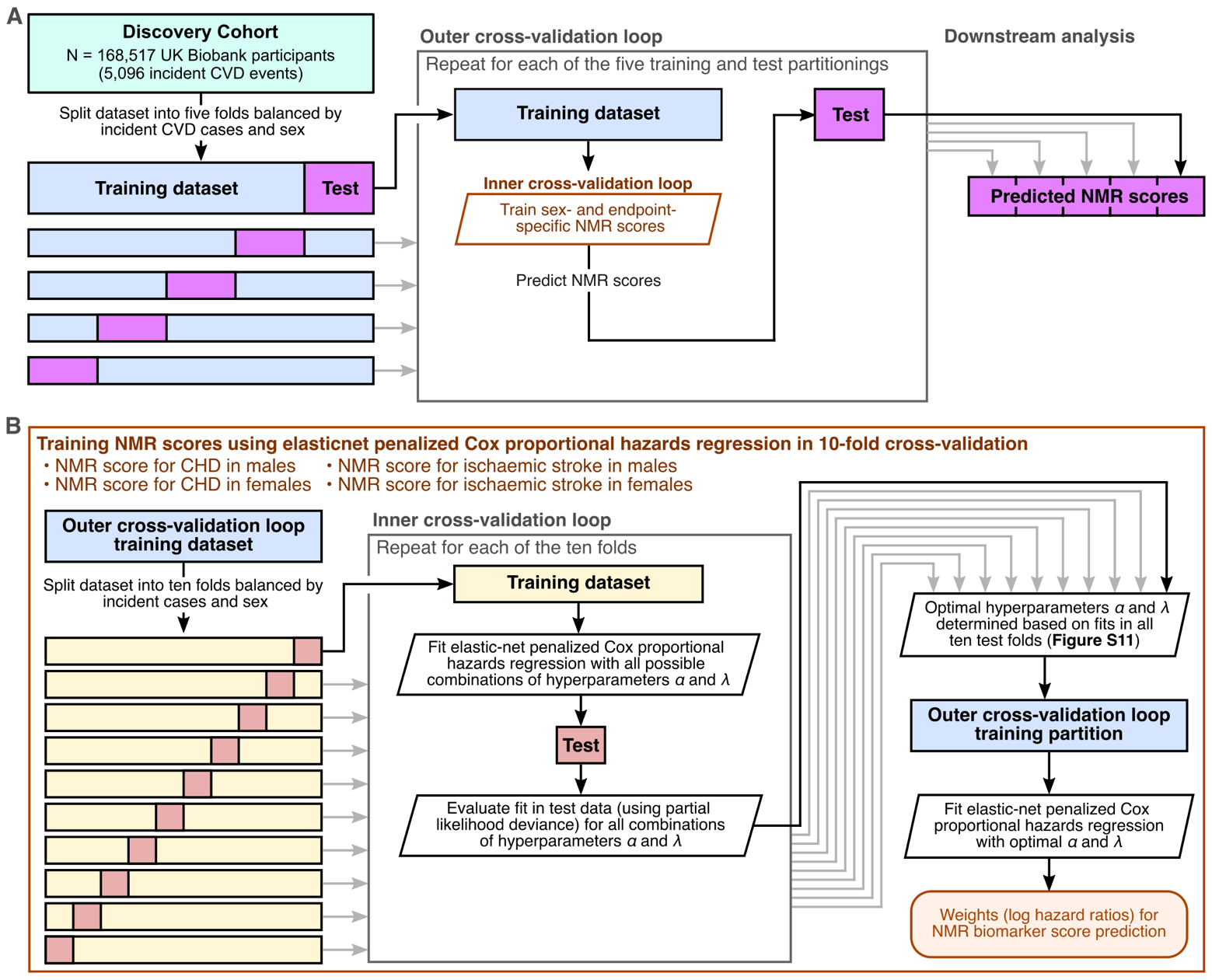

Figure S10: Schematic of NMR biomarker score training

**A)** Schematic showing the nested cross-validation procedure used to train sex- and endpoint- specific NMR biomarker scores. **B)** Detailed schematic of the inner cross-validation loop used for hyperparameter tuning in each of the five outer cross-validation loops (**Supplementary Methods**).

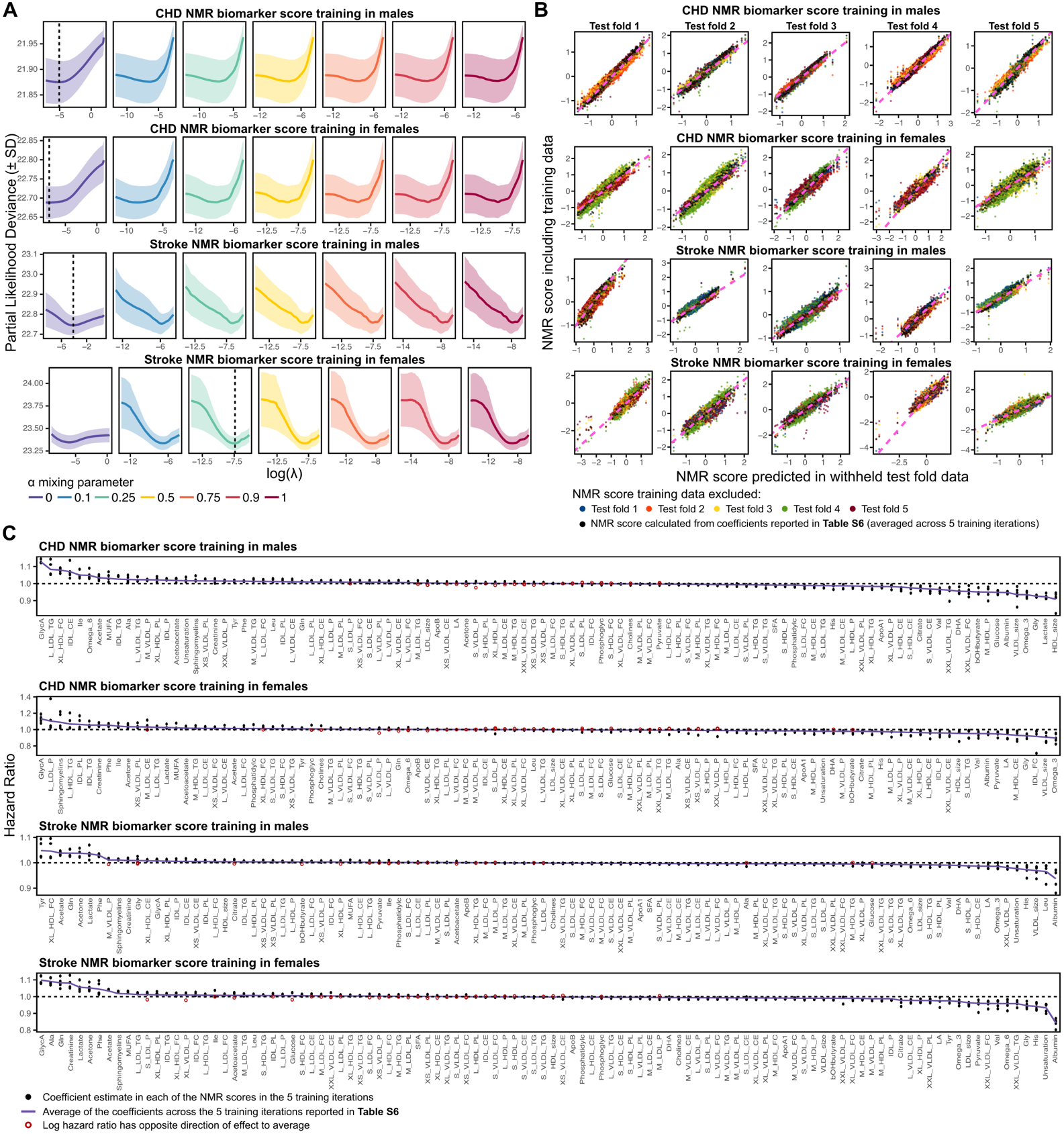

Figure S11: NMR biomarker score training

**A)** Selection of optimal NMR biomarker scores in one of the five training iterations (**Figure S10A**, **Supplementary Methods**). Each row of plots shows the grid-search for the optimal elasticnet parameters α and λ in 10-fold cross-validation (**Figure S10B**) for one of the four sex- and endpoint- specific NMR biomarker scores. At each tested α and λ the coloured line shows the mean of the partial likelihood deviance of the Cox proportional hazards regression for 10-year risk of the given endpoint across the 10 test folds. The coloured ribbon shows the standard deviation across the 10 test folds. The vertical black dashed line in each row of plots shows the optimal elastic-net fit; the combination of α and λ parameters that minimised the mean partial likelihood deviance. **B)** Concordance of NMR scores across the five training iterations when predicted in each of the five withheld test folds (**Figure S10A**). Each row of plots corresponds to one of the four sex- and endpoint- specific NMR scores, and each column corresponds to one of the five withheld test folds. Each scatterplot compares the NMR scores predicted in the withheld test fold (x-axes) to the NMR scores from the other four training iterations (y-axes), which included that test fold as part of their training dataset. Points on each plot are coloured to distinguish between the five training iterations. The black points show the NMR score computed from the average of coefficients across the five-training iterations, which we report in **Table S6** as the final representative weights for computing the NMR scores in new samples. The pink diagonal line show y=x, where points would fall if NMR scores from all training iterations were identical across samples. **C)** Compares the hazard ratios for NMR biomarkers for the optimal NMR scores in each of the five training iterations (black and red points). The purple line shows the average of the hazard ratio estimates reported in **Table S6**. Red points show instances where the direction of hazard ratios (<1, >1) differed between training iterations.

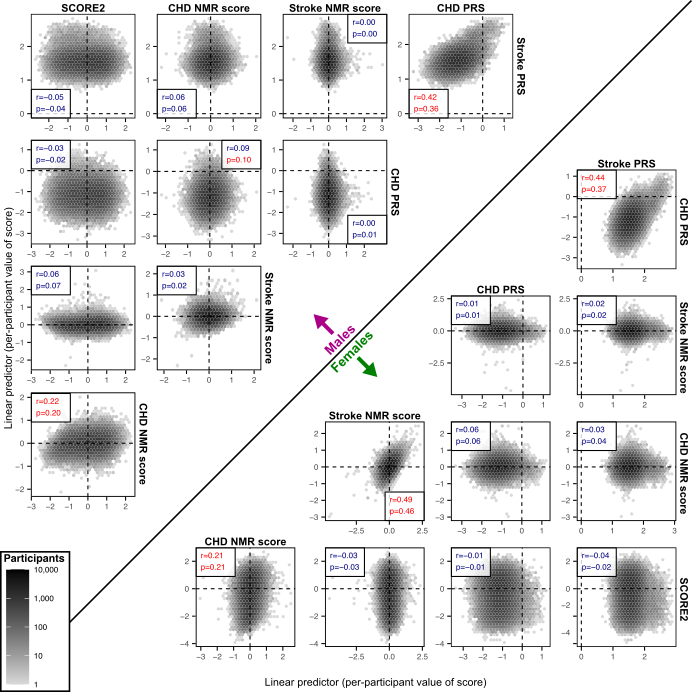

Figure S12: Pairwise correlation between SCORE2, NMR scores, and PRSs in males and females

Sex-stratified pairwise hexbin plots comparing densities of computed values (linear predictors) of SCORE2, the sex-specific CHD NMR score (**Table S6**), the sex-specific stroke NMR score (**Table S6**), the CHD PRS (PGS000018), and stroke PRS (PGS000039) in 168,517 UK Biobank participants included in the study discovery cohort (72,441 males, 96,076 females). Pearson’s (r) and Spearman’s (ρ) correlation coefficients are given in of each plot, blue where the magnitude < 0.1, and red where the magnitude was > 0.1.

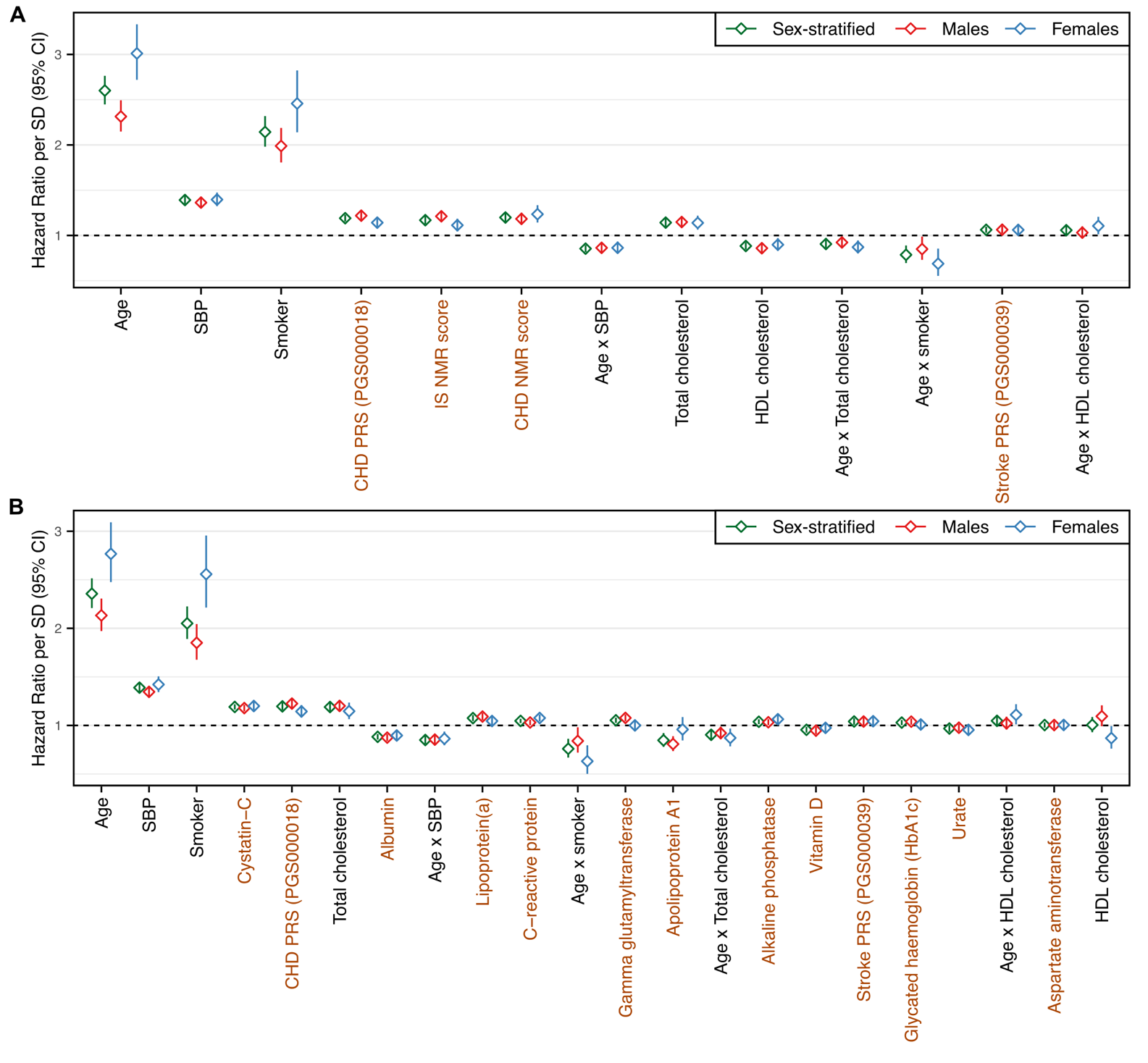

Figure S13: Independent associations of clinical biomarkers, NMR scores, PRS, and risk factors

**A)** Comparison of hazard ratios for 10-year CVD risk for NMR scores, PRSs and SCORE2 risk factors in sex-stratified and sex-specific (fit in males and females separately) multivariable Cox proportional hazards regression in the discovery cohort. Risk factors and age-interaction terms fit in the model are those comprising SCORE2 (**Supplementary** **Methods**). Details on the NMR scores are given in **Table S6** and the **Supplementary Methods**. Hazard ratios are per standard deviation increase in the respective NMR score, PRS, or risk factor, or per SD increase in age per SD increase in the respective risk factor. Predictors are arranged from left to right in ascending order of P-value in the sex-stratified analysis. Hazard Ratios, 95% CI, and P-values are detailed in **Table S21**. **B)** Comparison of hazard ratios for 10-year CVD risk for clinical biomarkers, PRSs, and SCORE2 risk factors. The 11 clinical biomarkers were those used in multivariable models (**Table S7**, **Methods**). Hazard Ratios, 95% CI, and P-values are detailed in **Table S21**.

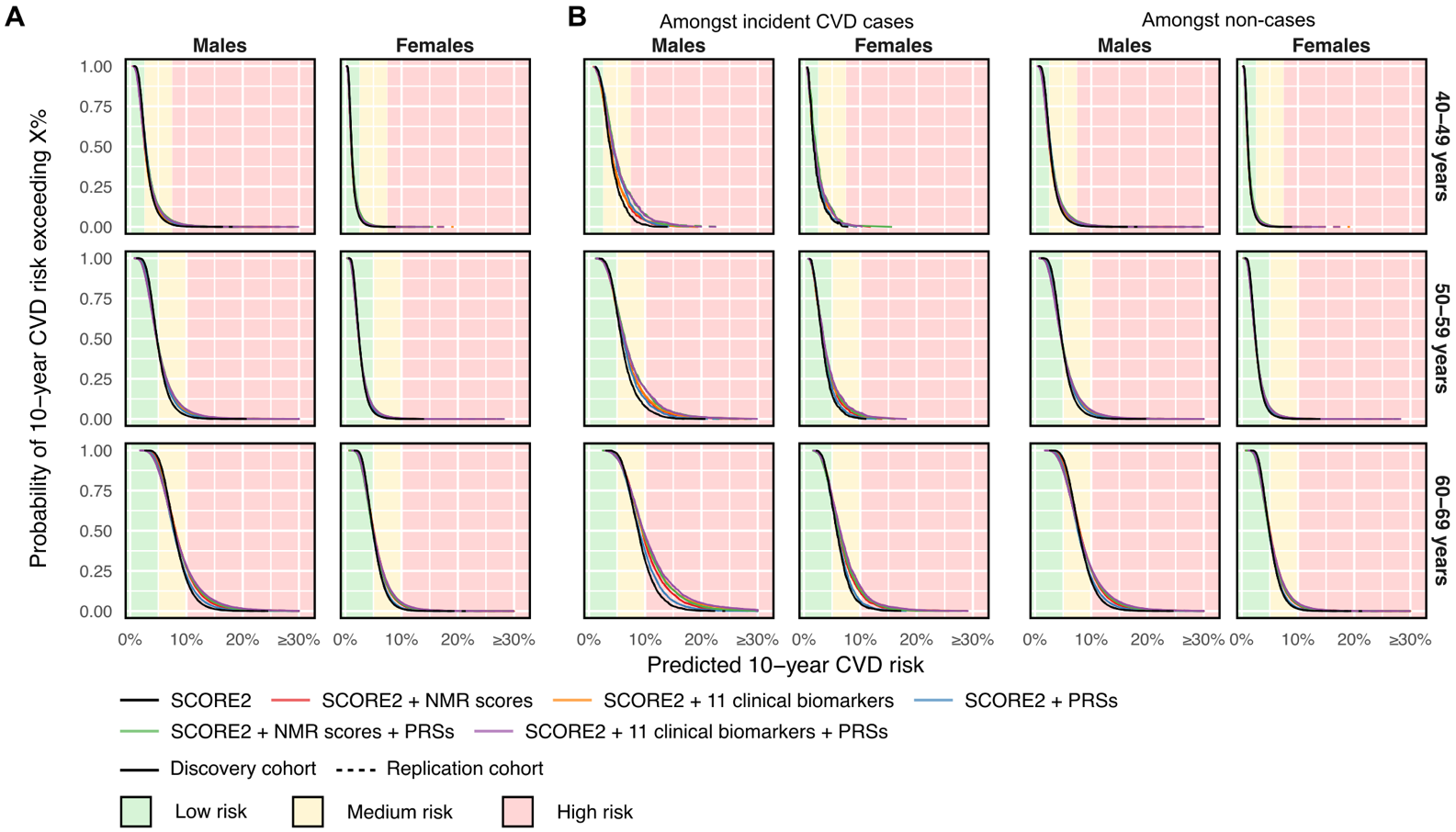

Figure S14: Comparison of distributions of predicted 10-year CVD risk when extending SCORE2

**A)** Probability of predicted 10-year CVD risk exceeding X% risk in the discovery and replication cohorts given a participant’s sex and decade of baseline age. **B)** Probability of predicted 10-year CVD risk exceeding X% risk further stratified amongst those with incident CVD and those without (non-cases). **A–B)** Probability curves were calculated as one minus the empirical cumulative distributive function in each cohort in each panel separately. The x-axis was constrained to show up to a maximum of 30% risk. Risk categories were determined using age-specific risk thresholds recommended by the ESC 2021 guidelines for CVD prevention. The low-risk category includes those 40–49 years of age with 10-year CVD risk <2.5% and those 50–69 years of age with 10-year CVD risk <5%. The medium-risk category includes those 40–49 years of age with 10-year CVD risk ≥2.5% and <7.5% and those 50–69 years of age with 10-year CVD risk ≥5% and <10%. The high-risk category includes those 40–49 years of age with 10-year CVD risk ≥7.5% and those 50–69 years of age with 10-year CVD risk ≥10%.

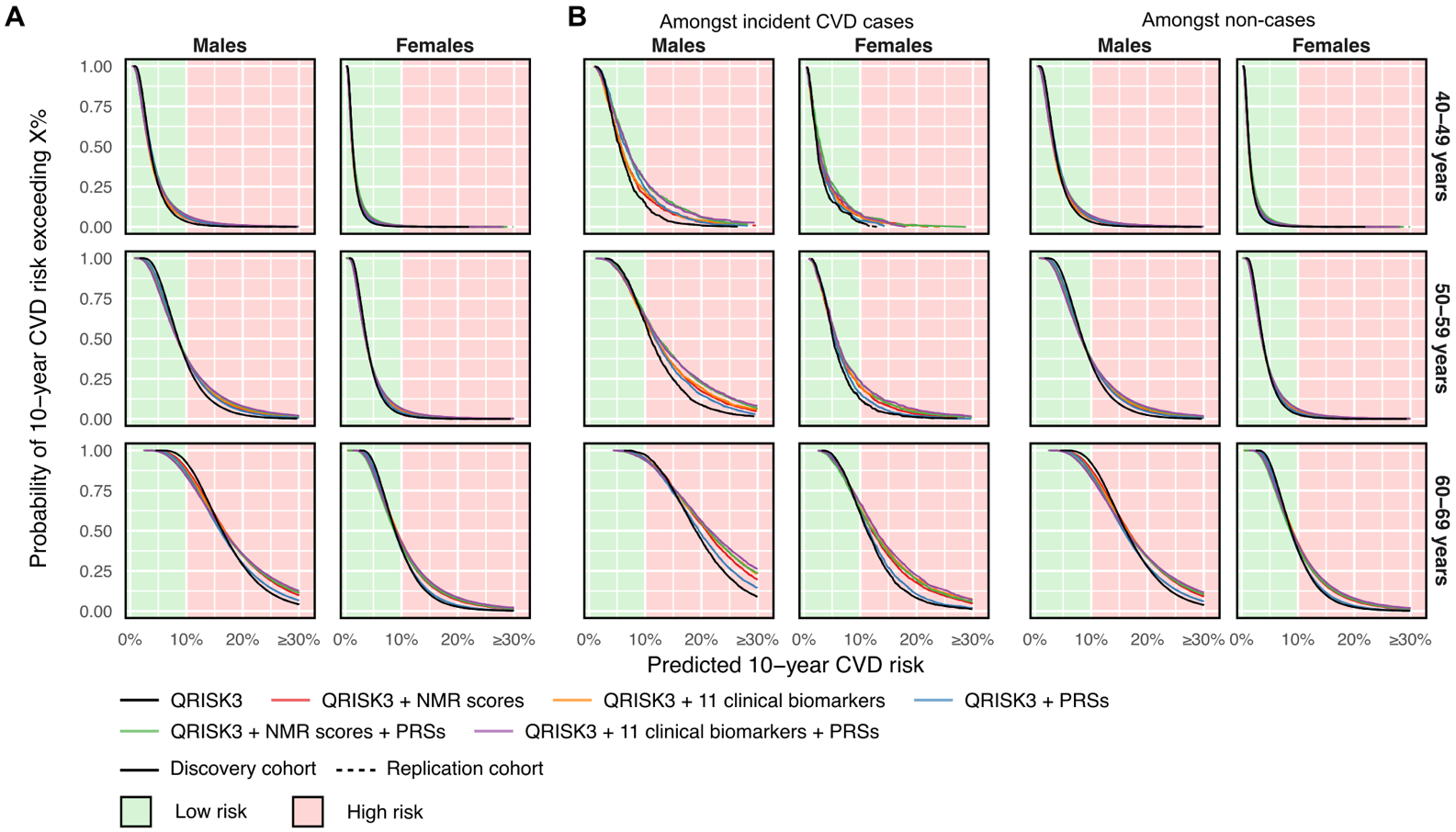

Figure S15: Comparison of distributions of predicted 10-year CVD risk when extending QRISK3

**A)** Probability of predicted 10-year CVD risk exceeding X% risk in the discovery and replication cohorts given a participant’s sex and decade of baseline age. **B)** Probability of predicted 10-year CVD risk exceeding X% risk further stratified amongst those with incident CVD and those without (non-cases). **A–B)** Probability curves were calculated as one minus the empirical cumulative distributive function in each cohort in each panel separately. The x-axis was constrained to show up to a maximum of 30% risk. Risk categories were determined using the 10% risk thresholds recommended by the UK’s National Institute for Health and Care Excellence (NICE) 2023 guidelines for CVD risk assessment and reduction.

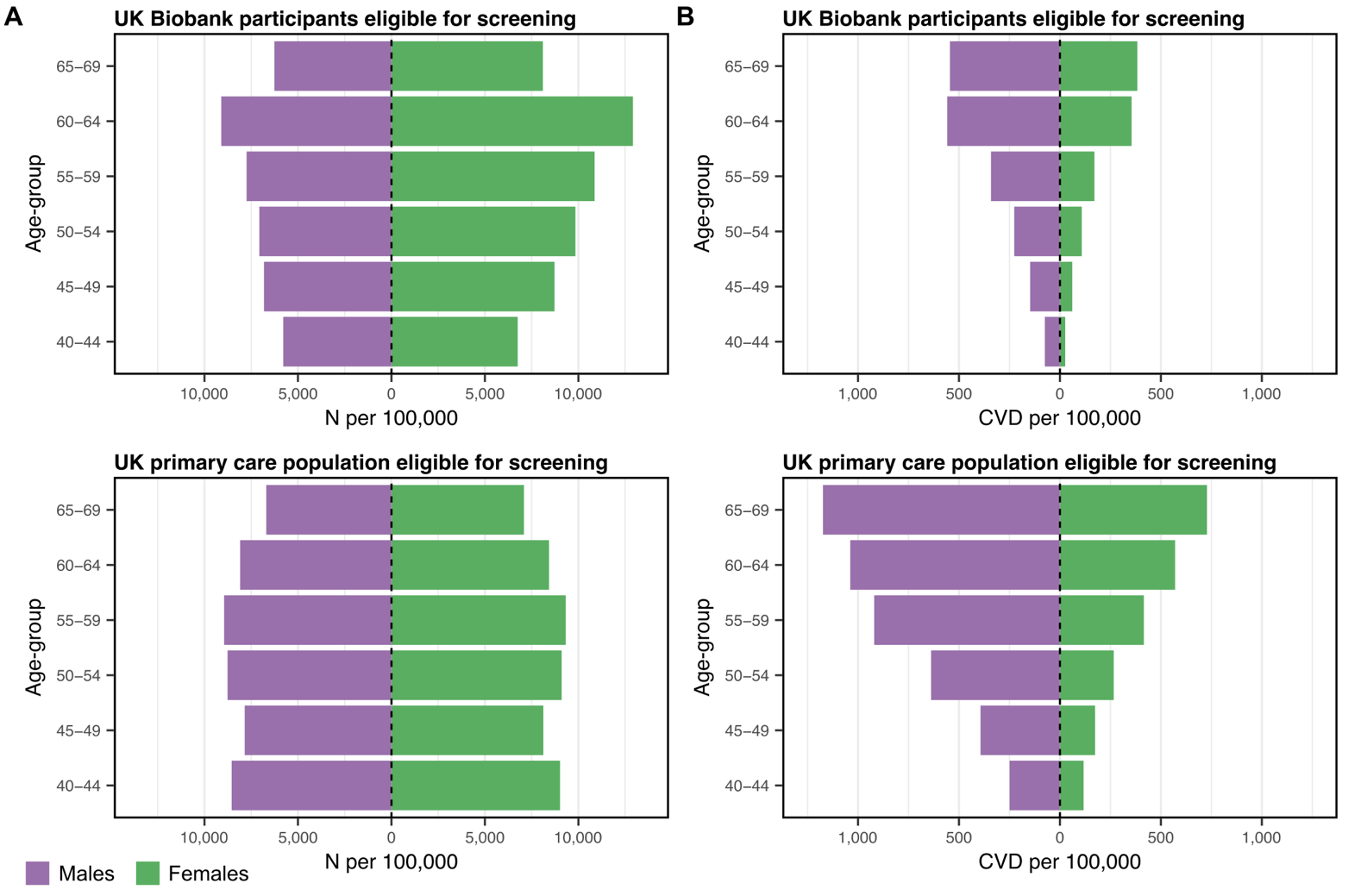

Figure S16: Demographic comparison of UK Biobank to the UK primary care population

**A)** Comparison of numbers per 100,000 in each age and sex group between the UK Biobank study population and the UK primary care population eligible for screening. Numbers per 100,000 in the UK primary care population eligible for screening were obtained from the mid-2023 population estimates in the UK published by the Office for National Statistics (**Supplementary Methods**). **B)** Comparison of incident CVD cases within 10-years per 100,000 in each age and sex group between the UK Biobank study population and the UK primary care population eligible for screening. CVD events per 100,000 in the UK primary care population eligible for screening were based on population sizes from the Office of National Statistics and 10-year CVD incident rates published by Sun *et al*. 2021 obtained from CVD- and statin- free primary care patients attending general practices between 2004 and 2017 (**Supplementary Methods**). **A–B)** Numbers and cases per 100,000 are detailed in **Table S22**.

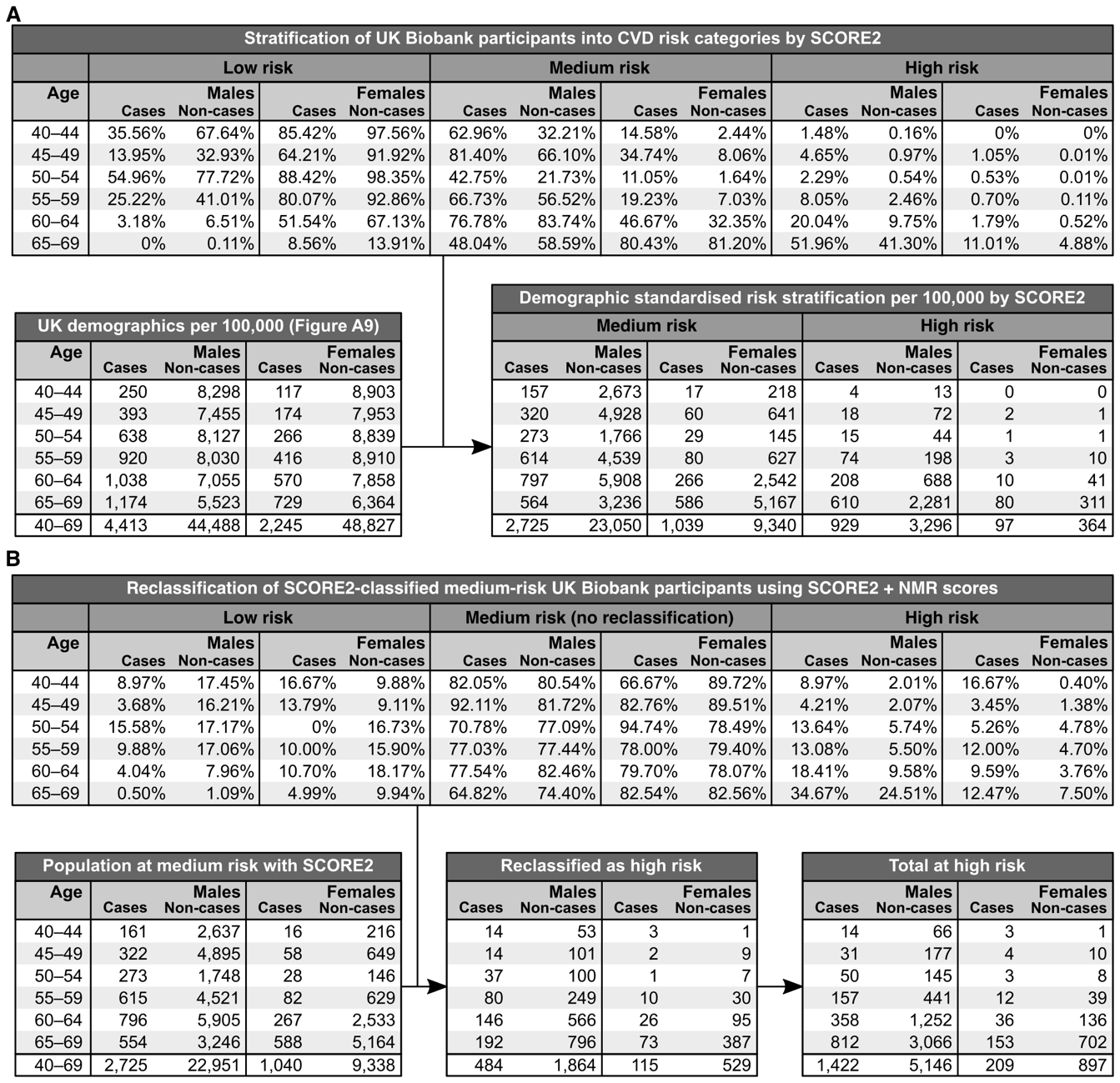

Figure S17: Schematic illustrating demographic standardisation of risk stratification

**A)** Standardisation of risk stratification by SCORE2 in the discovery cohort (n=168,517; 5,096 CVD cases) to the demographics and expected CVD incidents rates of the UK primary care population eligible for screening. The first table shows in the 168,517 UK Biobank participants, within each 5-year age-group and sex, the proportion of incident CVD cases and non-cases allocated to each risk category when using SCORE2 alone (**Methods**). Within each row, percentages sum to 100% with each sex and cases and non-cases separately. These proportions were then applied to the number of CVD cases and non-cases within 10-years per 100,000 estimated in the UK primary care population eligible for screening (**Figure S18**; **Supplementary Methods**) to obtain the demographic-standardised number of cases and non-cases allocated to each risk category by SCORE2. **B)** Targeted risk-reclassification with SCORE2 + NMR scores in the subset population classified as medium risk by SCORE2 alone. The first table shows in the 56,482 discovery cohort participants with complete data on NMR scores classified as medium-risk by SCORE2 alone, within each 5-year age-group and sex, the proportion of incident CVD cases and non-cases allocated to each risk category when SCORE2 + NMR scores (**Methods**). These proportions were then applied to the demographic-standardised number of cases and non-cases with complete data on NMR scores estimated to be at medium risk by SCORE2 alone to calculate the number predicted to be re-stratified to the high-risk category by SCORE2 + NMR scores.

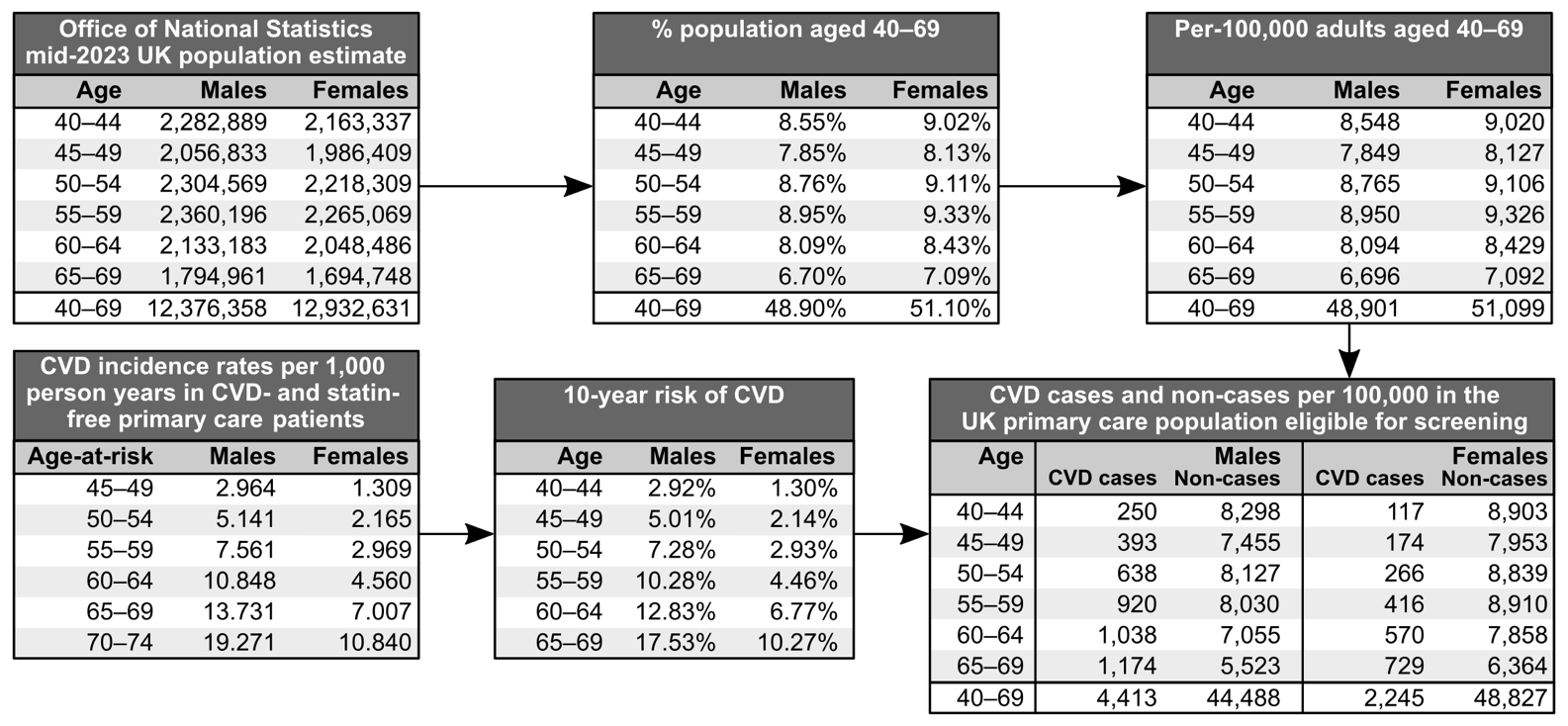

Figure S18: Derivation of demographic standardisation factors

for standardising risk stratification in the study cohort to the demographics and expected CVD incidents rates of the UK primary care population eligible for screening. Bottom right: details the number expected incident CVD cases and non-cases within 10-years per 100,000 estimated in the UK primary care population eligible for screening (**Supplementary Methods**) used in the standardisation procedure (**Figure S17**). Tables in the top row show how the number of males and females in each age-group were derived from the mid-2023 UK population estimates published by the Office for National Statistics. Tables on the bottom left show how CVD case numbers in males and females in each age-group were estimated using previously published estimates of per-1000-year CVD incidence obtained from primary care records of CVD- and statin- free patients^12^ attending general practices in the UK between 2004–2017. 10-year CVD risks were calculated from per-1000-year CVD incidence rates as 1 minus the exponential CVD-free survival in the 10-years ahead.

### Supplementary Tables

Table S1: Characteristics of discovery and replication cohorts

Discovery cohort: subset of UK Biobank study cohort with NMR metabolomics data available as of July 2023. Replication cohort: subset of remaining UK Biobank study cohort for whom early access to NMR metabolomics data were available from Q3 of 2024 onwards under UK Biobank project #30418. SBP: systolic blood pressure. HDL: High density lipoprotein. LDL: Low density lipoprotein. sd: standard deviations. IQR: interquartile range.

Table S2: NMR metabolite biomarker details.

Biomarker: short descriptive variable name used to label each column of biomarker data by the ukbnmr R package. Display name: short label used for plotting. Description: full name or description for the biomarker. UKB Field ID: field ID for the biomarker in UK Biobank. Units: units of absolute concentration. Type: whether the biomarker is composite biomarker (sum of two or more other biomarkers), a ratio, or a non-derived biomarker. The 106 non-derived biomarkers were used for NMR score training. Group: biological grouping of biomarkers. Sub-group: additional grouping for lipoprotein sub-fractions, fatty acids, and amino acids. % missing: % of samples in each analysis cohort (**Figure 1**) with missing data (missing at random; *e.g.,* due to technical error; **Supplementary Methods**) for that biomarker. Further details, such as derivation formulae for composite and derived biomarkers, can be found in the ukbnmr R package.

Table S3: Biochemistry assay biomarker details.

Biomarker: short descriptive name for biomarker. Description: full name or description for the biomarker. Classification: broad functional grouping of each biomarker. UKB Field ID: field ID for the biomarker in UK Biobank. Reportability Field ID: field ID in UK Biobank containing information on each missing data value, whether it was reported as missing due to (1) the biomarker concentration being above limits of reportability or detection, (2) the biomarker concentration being below limits of reportability or detection, or (3) some other reason. Units: units of absolute concentration as reported by UK Biobank. Analysis method: method of quantification as reported on the UK Biobank showcase for each biomarker. Analytical platform: machine used for biomarker quantification. Assay manufacturer: company which supplied the machine or diagnostic test. % missing: % of samples in each analysis cohort (**Figure 1**) with missing data (missing at random; *e.g.,* due to technical error; **Supplementary** **Methods**) for that biomarker. Note HDL and total cholesterol were not assessed for incremental improvement over SCORE2 as they were used in the calculation of SCORE2 (**Supplementary Methods**)**.**

Table S4: Incremental 10-year CVD risk discrimination over SCORE2 for individual biomarkers

Data underlying and extending **Figure 2**. Details the ΔC-index for 10-year risk of incident CVD for each of the 249 NMR biomarkers (**Table S2**) and 28 clinical biochemistry assays (**Table S3**) over SCORE2 alone in sex-stratified analyses in the discovery cohort (n=168,517; 5,096 CVD cases), replication cohort (n=128,946; 3,823 CVD cases), and in pooled analyses of the discovery and replication cohorts (n=297,463; 8,919 CVD cases). In each cohort a total of 277 SCORE2 + biomarker model fits were compared to SCORE2 alone (**Methods**). Technical details on the computation of standard errors, 95% confidence intervals, and p-values are reported in the **Supplementary Methods**. C-indices under the SCORE2 column headings were calculated in the subset of participants with measurements for the respective biomarker. Hazard ratios per standard deviation increase in the biomarker are reported for sex-stratified Cox proportional hazards regression fit in the discovery cohort with SCORE2 as an offset term. These hazard ratios were used to predict the SCORE2 + biomarker model fits when calculating C-indices and ΔC-indices in the replication and pooled discovery + replication analyses (**Methods**). FDR: false-discovery rate adjusted P-value across the 277 tested biomarkers in each cohort. Model fits are organised in ascending order of ΔC-index P-value in the pooled analysis of the discovery and replication cohorts.

Table S5: Risk discrimination sensitivity analyses for individual biomarkers

Extends **Table S4** to include sensitivity analyses when (1) analysing males and females separately, (2) using QRISK3 as the conventional risk score, and (3) using a narrower set of criteria for identifying incident CVD events (**Methods**). Risk score + biomarker model fits are ordered in ascending order of ΔC-index P-value in the main analysis, i.e. as in **Table S4**.

Table S6: Coefficients for computing NMR biomarker scores in new samples

Details the sex-specific coefficients for the coronary heart disease (CHD) and ischaemic stroke (IS) NMR scores obtained using the NMR score training procedure (**Figure S10**, **Supplementary Methods**) in the discovery cohort (n=168,517; 5,096 CVD cases). Coefficients weights are per standard deviation increase in the respective NMR biomarker. Sex-specific means and standard deviations for each NMR biomarker are reported. Provided NMR biomarkers have similar means and standard deviations in the new samples, NMR scores can be computed by standardising their concentrations in the respective sex, multiplying them by the reported coefficient, then summing together the result. The variance explained indicates the relative contribution of each biomarker to the respective NMR score (**Supplementary Methods**).

Table S7: Multivariable model fits for combining biomarkers, NMR scores, and PRSs with SCORE2

Data underlying and extending **Figure 3**. Details the sex-specific Cox proportional hazards regressions fit in the discovery cohort (n=168,517; 5,096 CVD cases) to create multivariable models of (1) SCORE2 + PRSs, (2) SCORE2 + NMR scores, (3) SCORE2 + clinical biomarkers, (4) SCORE2 + NMR scores + PRSs, and (5) SCORE2 + clinical biomarkers + PRSs. For each model, Cox proportional hazards regressions were fit for 10-year CVD using SCORE2 as an offset term. Hazard ratios are per standard deviation increase in the respective biomarker, NMR score, or PRS. Means and standard deviations in the subset of participants with complete data for each model are reported. Log hazard ratios were used to predict the sex-specific models in the replication cohort (**Supplementary Methods**) and can be used to compute models in new samples after standardisation. Biomarkers in models including clinical biomarkers were those that had FDR significant ΔC-index over SCORE2 alone when testing individual biomarkers (**Table S4**). Details on the NMR scores are given in **Table S6** and the **Supplementary Methods**. CHD: coronary heart disease. IS: ischaemic stroke.

Table S8: Incremental risk discrimination for NMR scores, combined biomarkers, and PRSs

Data underlying and extending **Figure 4A**. Details the ΔC-index for 10-year risk of incident CVD for each multivariable model over SCORE2 alone in sex-stratified analyses in the discovery cohort (n=168,517; 5,096 CVD cases), replication cohort (n=128,946; 3,823 CVD cases), and in pooled analyses of the discovery and replication cohorts (n=297,463; 8,919 CVD cases). For each model, a sex-stratified Cox proportional hazards regression was fit in the discovery cohort with SCORE2 as an offset term (**Table S7**), then model fits were predicted in each cohort when calculating C-indices and ΔC-indices (**Supplementary Methods**). For models including clinical biomarkers or NMR scores, the number of participants with complete data on the underlying 11 clinical biochemistry biomarkers (**Table S7**) or 106 NMR biomarkers (**Table S6**) are reported.

Table S9: Risk discrimination sensitivity analyses for NMR scores, combined biomarkers, and PRSs

Extends **Table S8** to include sensitivity analyses when (1) analysing males and females separately, (2) using QRISK3 as the conventional risk score, and (3) using a narrower set of criteria for identifying incident CVD events (**Methods**).

Table S10: Categorical net reclassification for NMR scores, combined biomarkers, and PRSs

Data underlying and extending **Figure 4B**. Details the categorical net reclassification improvement (NRI) analysis assessing the incremental value of the five multivariable models over SCORE2 for stratifying individuals into low, medium, and high-risk groups based on ESC 2021 recommended risk thresholds for treatment consideration (**Methods**). Results are shown for categorical NRI analyses run in the discovery cohort (n=168,517; 5,096 CVD cases), replication cohort (n=128,946; 3,823 CVD cases), and in pooled analyses of the discovery and replication cohorts (n=297,463; 8,919 CVD cases) separately. CVD cases were correctly reclassified if, based on their predicted absolute risk, they were moved from a lower risk group to higher risk group with the multivariable model compared to SCORE2. Non cases were correctly reclassified if, based on their predicted absolute risk, they were moved from a higher risk group to lower risk group with the multivariable model compared to SCORE2. Standard errors, 95% confidence intervals, and P-values were estimated in a bootstrap procedure with 1,000 bootstraps (**Supplementary Methods**). Numbers allocated to each risk category by each model are detailed in **Table S11**.

Table S11: Risk stratification and reclassification for NMR scores, combined biomarkers, and PRSs

Data underlying **Table S10**. Details the number of CVD cases and non-cases allocated to each possible combination of risk groups when comparing SCORE2 to each of the five multivariable models adding clinical biomarkers, NMR scores, and/or PRSs to SCORE2. Risk categories were defined using age-group specific risk thresholds recommended by the ESC 2021 guidelines for clinical decision making on initiating risk factor treatment for CVD prevention (**Methods**).

Table S12: Categorical net reclassification sensitivity analyses

Extends **Table S10** to include sensitivity analyses when (1) analysing males and females separately, (2) using QRISK3 as the conventional risk score, and (3) using a narrower set of criteria for identifying incident CVD events (**Methods**). Risk categories were defined using ESC 2021 guideline age-group specific risk thresholds when using SCORE2 as the conventional risk score (**Methods**) or using the NICE 2023 guideline recommend risk threshold (10%) when using QRISK3 as the conventional risk score (**Supplementary Methods**).

Table S13: Risk stratification and reclassification in sensitivity analyses

Data underlying **Table S12**. Details the number of CVD cases and non-cases allocated to each possible combination of risk groups in the categorical net reclassification sensitivity analyses (**Methods**). Risk categories were defined using ESC 2021 guideline age-group specific risk thresholds when using SCORE2 as the conventional risk score (**Methods**), or using the NICE 2023 guideline recommend risk threshold (10%) when using QRISK3 as the conventional risk score (**Supplementary Methods**).

Table S14: Proportions classified as high risk by SCORE2 and QRISK3 in different age-groups

Numbers of males and females in each five-year age group classified as high-risk when using either SCORE2 or QRISK3 and risk thresholds recommended by the ESC 2021 and NICE 2023 guidelines respectively. ESC 2021 guidelines recommend age-specific risk thresholds on SCORE2 (≥7.5% risk and ≥10% risk for people under 50 and 50 or older respectively), whilst NICE 2023 guidelines recommend a 10% risk threshold on QRISK3.

Table S15 Incremental value for CVD prevention per 100,000 in the UK eligible for screening

Data underlying and extending **Figure 5**. Details the incremental value for primary prevention of CVD per 100,000 in the UK eligible for screening when adding NMR scores, clinical biomarkers, and/or PRSs to SCORE2. Two screening strategies were assessed: (1) population-wide screening, in which all people eligible for screening were assessed with each model; and (2) targeted screening, in which people were first stratified into low, medium, and high-risk groups using SCORE2 alone, then those allocated to the medium risk group were re-assessed using the models adding NMR scores, clinical biomarkers, and/or PRSs to SCORE2. Estimates were derived by standardising risk stratification in the respective study cohort to the demographics and expected CVD incidents rates (6,686 per 100,000) of the UK primary care population eligible for screening (**Methods**). Standard errors, 95% confidence interval, and P-values were estimated via a bootstrap sampling procedure with 1000 bootstraps (**Supplementary Methods**). Differences in statistics under the SCORE2 column headings between models reflect differences in data availability for each pair of model comparisons similar to other analyses.

Table S16: Incremental value for CVD prevention per 100,000 in sensitivity analyses

Extends **Table S15** to include sensitivity analyses when (1) analysing males and females separately, (2) using QRISK3 as the conventional risk score, and (3) using a narrower set of criteria for identifying incident CVD events (**Methods**). Risk categories were defined using ESC 2021 guideline risk thresholds when using SCORE2 as the conventional risk score (**Methods**) or using the NICE 2023 guideline recommend risk threshold when using QRISK3 as the conventional risk score (**Supplementary Methods**).

Table S17: Definition of established atherosclerotic cardiovascular disease.

List of UK Biobank fields, record sources, and corresponding field codes used to identify participants with established atherosclerotic cardiovascular disease (ASCVD) to exclude from the study based on the ESC 2021 guidelines on cardiovascular disease prevention. ICD-10: International Classification of Diseases (ICD) 10^th^ edition. ICD-9: ICD 9^th^ edition. OPCS-4: Office of Population Census and Surveys (OPCS) Classification of Interventions and Procedures version 4. OPCS-3: OPCS Classification of Interventions and Procedures version 3. ICD-9 codes are present in the hospital episode statistics up until 30^th^ March 1996 for hospital events in Scotland and up to 5^th^ February 1995 for hospital events in Wales. All hospital events in England are coded with ICD-10 codes. OPCS-3 codes are present in the hospital episode statistics up to 30^th^ December 1988 for operations and procedures performed in Scotland.

Table S18: List of prescription medications used for sample exclusion criteria

List of prescription medications used to define the statins and lipid lowering medication sample exclusion criteria. Medication codes and labels are as given in UK Biobank field #20003. Lipid lowering medications qualifying for the exclusion criteria were determined by cross-referencing the list of medications present in UK Biobank with the British National Formulary (BNF) chapter 2.12: Lipid-regulating drugs with the restriction that the drug indications must include lipid-lowering for prevention of cardiovascular diseases. Medications in the BNF chapter 2.12 whose lipid-lowering was a side-effect for a drug not intended for prevention of cardiovascular diseases were not included in the sample exclusion criteria.

Table S19: List of prescription medications used when calculating QRISK3

List of prescription medications used when identifying participants with erectile dysfunction, or taking blood pressure lowering medications, systemic corticosteroids, or atypical antipsychotics. Medication codes and labels are as given in UK Biobank field #20003. Medications in each category were determined by cross-referencing the list of medications present in UK Biobank with relevant British National Formulary chapters and indication data, as well as relevant lists of medications in the QRISK3 publication. See **Supplementary Methods** for further details.

Table S20: Cohort characteristics for incident CHD and ischaemic stroke

**A)** Characteristics 10-year follow-up for incident CHD in the 168,517 UK biobank participants in the study discovery cohort. **B)** Characteristics of 10-year follow-up for incident ischaemic stroke in the 168,517 UK biobank participants included in the study discovery cohort. sd: standard deviations. IQR: interquartile range.

Table S21: Multivariable model fits of NMR scores, clinical biomarkers and PRSs with risk factors

Details Cox proportional hazards regression fit in the discovery cohort (n=168,517; 5,096 CVD cases) for (1) SCORE2 risk factors + NMR scores + PRSs, (2) SCORE2 risk factors + clinical biomarkers + PRSs, (3) QRISK3 risk factors + NMR scores + PRSs, and (4) QRISK3 risk factors + clinical biomarkers + PRSs. Details on the NMR scores are given in **Table S6** and the **Supplementary Methods**. The 11 clinical biomarkers were those used in multivariable models (**Table S7**, **Methods**). For continuous measures, hazard ratios are per standard deviation increase. For QRISK3, hazard ratios for different smoking statuses are relative to non-smokers, and hazard ratios for different ethnicities are relative to those self-reporting White ethnicity or not stating their ethnicity. Details on QRISK3 risk factors are provided in the **Supplementary Methods**. Age P1 and Age P2 correspond to sex-specific polynomial terms used by QRISK3 to model non-linear effects of age. In females, Age P1 is Age^–2^ and Age P2 is Age. In males, Age P1 is Age^–1^ and Age P2 is Age^3^. BMI P1 and BMI P2 correspond BMI^–2^ and BMI^–2^ × log(BMI) respectively. Note that the large number of variables and interaction terms used by QRISK3 meant that Cox proportional hazards regressions did not converge, and some hazard ratios for some coefficients are either missing or have extreme implausible values and 95% confidence intervals.

Table S22: Demographic comparison of UK Biobank to the UK primary care population

Data underlying and extending **Figure S16**. Comparison of numbers and incident CVD events per 100,000 in each age and sex group between the UK Biobank study populations and the UK primary care population eligible for screening (**Methods**). Numbers and CVD events per 100,000 in the UK primary care population eligible for screening are estimates (**Supplementary Methods**): Numbers per 100,000 were obtained from the mid-2023 population estimates in the UK published by the Office for National Statistics, and CVD events per 100,000 were based on these population sizes multiplied by 10-year CVD incident rates published by Sun *et al*. 2021 obtained from CVD- and statin- free primary care patients attending general practices between 2004 and 2017.
